# Trends and Characteristics during 17 Years of Naloxone Distribution and Administration through a Harm Reduction Program in Pittsburgh, Pennsylvania

**DOI:** 10.1101/2024.11.21.24317731

**Authors:** Nabarun Dasgupta, Alice Bell, Malcolm Visnich, Maya Doe-Simkins, Eliza Wheeler, Adams L. Sibley, Maryalice Nocera, Amy E. Seitz, Dorothy Chang, Summer Barlow, Zachary Dezman, Jana McAninch

## Abstract

**Objective:** Describe time trends during 17.6 years of community-based naloxone distribution.

**Methods:** Analysis of administrative records from a harm reduction program in Pittsburgh, Pennsylvania, USA, collected during encounters for overdose education, naloxone dispensing and refills. Monthly time trends were analyzed using segmented regression. Programmatic context aided interpretation of quantitative findings. We also evaluated impacts of 2014 state legislation loosening naloxone prescribing requirements and providing Good Samaritan protections.

**Results:** From July 2005 to January 2023 there were 16,904 service encounters by 7,582 unique participants, resulting in 70,234 naloxone doses dispensed, with 5,521 overdose response events (OREs), utilizing 8,756 naloxone doses. After legislation, new participants increased from 10.4 to 65.9 per month. New participants tended to be older (46 vs. 37 years), female (58% to 35%), White race, and more likely to be family/friends as opposed to people who use drugs themselves. Consequently, ORE per participant fell from 1.46 to 0.47 in the year after enactment. On average, 1.63 (95% CI: 1.60, 1.65) naloxone doses were administered per ORE, which did not change substantially over 17 years (*χ^2^*=0.28, 3 df, *p*=0.60) during evolution from prescription opioids, to heroin, to illicitly manufactured fentanyl. In 98.0% of OREs the person who experienced overdose “was okay”, i.e., survived. Emergency medical services were called in 16% of OREs overall, but <7% since 2019. There were 106 more emesis events per 1,000 OREs with 4mg nasal spray compared to intramuscular injection; and 48 per 1,000 more reports of anger. Titration of intramuscular naloxone was associated with lower rates of adverse events.

**Conclusions:** While state legislation created the environment for expansion, reaching previously underserved communities required intentional new programmatic development and outreach. Long-term consistency of <2 doses per ORE, high survival rate, and robust utilization all lend confidence in prioritizing naloxone distribution directly to people who use drugs and their social networks.

## INTRODUCTION

In the United States (US), fatal overdose rates have increased over the past four decades. The characteristics have changed from heroin dominance in the early 1990s to prescription opioids in the late 1990s and 2000s, to heroin again around 2013 [1,2]. By 2015, the emergence of non-pharmaceutical fentanyl and analogues played the most prominent role in fatal overdoses, and currently stimulant-opioid fatal overdoses are increasing [3,4]. After many years of unrelenting increases, the latest national data appear to show a 3% decline in annual overdose deaths in 2023 [5,6]. However, at the state level the results are heterogeneous: some states posted 15% reductions, and others showed marked increases.

Community-based distribution of the opioid overdose reversal agent, naloxone, has expanded considerably since federal funding support in 2018 and is a critical strategy for preventing fatalities [7]. Harm reduction programs train people who use drugs to recognize and respond to opioid-induced respiratory depression. People who access these services, by the nature of their ongoing drug use, are at the highest risk of overdose [8], yet there is a paucity of quantitative data characterizing peer reversal behaviors, particularly as the drug supply landscape has evolved. Specifically, in the context of community-based naloxone distribution, there are few published studies detailing time trends of adverse events, formulation effects, titration practices, changes during the COVID-19 pandemic, and infiltration of xylazine in the unregulated drug supply. Over the last two decades, state laws have also evolved to reduce barriers to community-based naloxone distribution. In the scientific literature, policy analyses of state laws have lent support for this continued practice [9], localized evaluations have been published [10–13], as have evaluations of barriers [14,15]. What has been not been adequately described is the longitudinal evolution of harm reduction programs themselves. As harm reduction programs become enduring and institutionalized with increased public health investment, it is imperative to understand long-term programmatic trajectories, which can, in turn, inform future policy decisions.

Specifically, there is limited documentation of how harm reduction programs adapt to changing drug supply, laws, pharmaceutical formulations, and societal norms over an extended time. Herein we describe trends and experiences over 17 years from one of the longest continuously operating overdose prevention programs in the world, at Prevention Point Pittsburgh, a harm reduction and syringe services program. With continuous data collection [16], this setting offers a unique opportunity to understand long-term time trends in both community practice and individual responses simultaneously.

Applying the Evidence-Making Intervention framework [17], this comprehensive manuscript balances programmatic context with quantitative findings. The authors come from three professional domains: harm reduction program staff, government and academic scientists, and public health advocates. Some of the authors were the earliest innovators and implementors of community-based naloxone distribution in the US [18–21]. Of note, Prevention Point Pittsburgh has maintained institutional memory via staff retention; one co-author (AB) has been directing the program in Pittsburgh for the entirety of the observation period and provided invaluable canonical information on changes in service delivery. The length of the manuscript reflects aspects that are of relevance to each of the three professional domains, and we encourage readers to make use of section headings to follow the storyline that is of greatest interest.

### About Naloxone

Naloxone is a mu-opioid receptor antagonist that displaces or prevents binding of opioid agonists such as heroin, fentanyl, and morphine. It was first synthesized in 1961 by Jack Fishman and Harold Blumberg. Naloxone was approved as a human prescription medication by the US Food and Drug Administration (FDA) in 1971 to treat opioid-induced respiratory depression (“overdose”), and is on the World Health Organization’s List of Essential Medicines [22]. It reverses opioid-induced respiratory depression rapidly but may also precipitate withdrawal in people who have opioid tolerance. Community-based naloxone distribution has become widely accepted in the US as a means of secondary prevention of overdose deaths, albeit with heterogeneity in enabling state governmental laws and policies [23].

### Brief History of Community Naloxone Distribution

Prevention Point Pittsburgh started distributing naloxone in 2005, after being inspired by formative work by the Chicago Recovery Alliance. Previously naloxone had been used exclusively in hospitals for managing anesthesia and by pre-hospital emergency medical service providers to reverse opioid overdose. In 1996, fueled by rising fatal heroin overdose among participants and staff, the Chicago Recovery Alliance [24] began distributing naloxone via their syringe services program to people who use drugs and their immediate social networks, an innovation marking the first known formal overdose education and naloxone distribution program in the world [25,26].

For the first decade of operations, the naloxone distribution program at Prevention Point Pittsburgh operated within a broader national context, which evolved from an environment of little support to codified scientific and legal protections. Given limited funding during the first 18 years (1996 to 2014) of broader intervention evolution, the development and implementation of new naloxone distribution initiatives within syringe services programs nationally was primarily through peer-based mentoring and technical assistance between programs. This was the case with Prevention Point Pittsburgh. Naloxone was purchased using smaller value unrestricted funds from sources such as t-shirt sales and donations to memorial funds from families who had lost a loved one to overdose. The staff time and cost to implement and deliver the services was absorbed by syringe service programs, viewed as an ethical imperative regardless of funding.

Using this unfunded interorganizational mentoring model, there were 48 programs in the US by 2010 [18], and 140 by 2014 [27]. Because naloxone was a prescription medication, these initiatives existed in a medico-legal gray area that generated onerous requirements on harm reduction programs. For example, from 2005 to 2014, a documented in-person medical encounter and individual prescription from a physician was required for Prevention Point Pittsburgh to dispense naloxone to a participant. After coordinated national advocacy by public health organizations, state level legislation, and accumulating scientific evidence, policies supporting naloxone distribution were established starting around 2014 nationally, and directly contributed to the expansion of the naloxone distribution initiative in Pittsburgh.

The advent of federal support for naloxone distribution also had an impact on Prevention Point Pittsburgh by creating an expanded community of harm reduction practice for innovation, diffusion, and communication. In 2014, a memo from the Substance Abuse and Mental Health Services Administration (SAMHSA) to the National Association of State and Territorial AIDS Directors (NASTAD) clarified that using federal funds for naloxone was an acceptable expenditure for state block grants [28]. The first new federal funding that explicitly allowed for naloxone distribution was the Health Resources and Services Administration (HRSA) 2015 Rural Opioid Overdose Reversal grant program [29]. Prior to federal funding, local governments and harm reduction programs used local and philanthropic funds to support naloxone distribution in Massachusetts [20], New York [14], New Mexico [30], San Francisco [31], Rhode Island [32], North Carolina [21], Baltimore [33], and Pittsburgh [19]. Prevention Point Pittsburgh operated within this community of practice, the activity of which centered around the listserv and monthly meetings of the Opioid Safety and Naloxone Network, facilitated for over a decade by co-author AB [34,35].

By the end of 2015, several key events paved the way for further development of Prevention Point Pittsburgh’s naloxone distribution program. Research emerged confirming that naloxone distribution via syringe services programs was effective at reducing overdose mortality [36] and was cost-effective [37,38]. Laws were passed in 43 states to support expansion [39,40]. Two new branded naloxone products (nasal spray and auto-injector) were approved for prescription use among lay persons [41] and heavily promoted by pharmaceutical manufacturers [42]. Harm reduction programs also created a Buyers Club to obtain low cost injectable naloxone directly from a different manufacturer [34]. The FDA supported development of nonprescription naloxone formulations by conducting studies of labeling instructions [43] and expediting review of new products [44]. Of direct relevance to Prevention Point Pittsburgh, Pennsylvania Act 139 was enacted on November 30, 2014, allowing standing orders and third-party naloxone prescriptions. Prevention Point Pittsburgh’s Medical Director issued a standing order for the organization, enabling naloxone distribution without requiring individual prescriptions. Other relevant contextual dates are cataloged in Supplemental Material Table S1.

### Research Questions

Five research questions were specified in the public pre-registration [45].

1. Did the utilization rate of naloxone and demographics of participants change after enabling state legislation was enacted?
2. After enactment of state legislation, what actions did the program take to focus uptake of naloxone directly to networks of people who use drugs?
3. Were program adaptations (e.g., site expansion) effective in improving naloxone uptake among communities of color in Pittsburgh?
4. Has the average number of doses of naloxone administered during an overdose response event changed over time as the drug supply has changed? Specifically, was more naloxone needed for reversing overdoses during the era of illicitly manufactured fentanyl, compared to previous periods where overdoses were due to heroin [46]?
5. Is the number of doses administered per overdose response event impacted by type of naloxone formulation?

Three additional questions were developed by the co-authors during the iterative analysis process and evaluated in accordance with the Evidence-Making Intervention (EMI) framework (described in Methods).

1. Did enactment of the Pennsylvania “Good Samaritan” law impact the proportion of overdoses response events in which 911 was called?
2. What were the circumstances of deaths reported after administration of naloxone?
3. Did adverse events differ by formulation of naloxone? And did titration of naloxone have an impact on adverse event rates?

## METHODS

### Conceptual Framework

This study was based on the EMI framework [17,47]. This framework shifts the locus of evidence production away from universally generalizable knowledge, which is common in traditional biomedical research. Instead, EMI prioritizes a more contextualized scientific process in which data and conclusions are generated through localized public health interventions serving immediate, applied needs. Therefore, the purpose of this analysis is not to present the hypothetically universal experience of naloxone distribution, but rather to examine one location in-depth to understand the forces that directly impacted service delivery and naloxone utilization. The application of the framework to the current investigation can be summarized using the six central tenets of EMI. In applying these principles in the Results section, “Programmatic Context” follows “Quantitative Results” for each set of variables analyzed.

1. *Material-discursive Process*: Naloxone distribution in Pittsburgh is not expected to be the same as anywhere else, yet there is value in understanding the local context. State policies and local drug supply considerations are made when interpreting quantitative data.
2. *Emergent, Contingent, Multiple effects:* Applied to this study, participant behaviors were expected to change over time. Overdose response practices naturally evolved over a 17-year period, instead of assumed to be static, as in shorter studies.
3. *Practice-based Matter-of-concern:* Of central relevance is how the concept of naloxone distribution was interpreted by program staff and locally adapted. For example, the program adapted to the COVID pandemic, and as new naloxone products and street drugs shifted. Therefore, contemporaneous contextual details are provided allow quantitative data to be interpreted with fidelity.
4. *Practice of Implementation:* How the intervention was delivered is of equal importance to other outcomes (e.g., biomedical or pharmacological). Therefore, logistical considerations and site expansion rationales are provided in detail, especially in ways that impacted participant recruitment and training of participants, and ultimately, the quantitative data.
5. *Performative Work of Science:* Administrative data were collected first and foremost for service delivery, and the scientific knowledge generated from their review is an added benefit. While data were collected with the intention of analysis, the questions asked of participants were also designed to gather information on reversals that would reveal opportunities for counselling and behavior change at the point of care.
6. *Equality of Knowledge:* Program staff’s experience of service delivery is of equal explanatory value as quantification of administrative records. Program staff were included in each step of the analysis process, and their experiences are recorded in the Results section, and they are co-authors of this manuscript.

The Equality of Knowledge principle, a recursive process for knowledge generation was applied, starting with whole-team generation of the research questions. The data analyst (ND) generated tabular and graphical representations of time trends for batches of variables. The team then assembled to discuss patterns, aberrations, policy impacts, public health implications, and topics for further investigation, including new research questions based on discussions of programmatic context. After the initial discussion, the analyst would prepare follow-up tables, developing statistical methods as the inquiry warranted, and refine time trend graphs, which were then presented at the following meeting. This recursive process was applied to each set of variables in the dataset until all variables had been analyzed and discussed. In addition to the five research questions elaborated in the pre-registration, the recursive process resulted in three additional research questions described above.

### Data Source

We analyzed naloxone dispensing records and participant intake forms from a multi-site comprehensive harm reduction program (e.g., syringe services provider) in Pittsburgh, Pennsylvania, US. Datasets were anonymized by Prevention Point Pittsburgh prior to analysis. Data were generated at either initial training encounters or refill requests by participants, the latter of which included questions about the overdose that the naloxone had been used during. Interviewers received training to ensure standardized data collection, and ongoing data quality assessments were conducted. These administrative records span July 24, 2005 to January 24, 2023; initial naloxone distribution started in late July 2005 and the first reversal was reported in August. Data were collected on standardized paper forms, with weekly manual data entry using an electronic record system. Keystroke entry and missing corrections for early years of data were necessary to standardize dates and syntactical conventions (e.g., comma versus semi-colon for list delimiting) using natural language processing (described in Supplemental Material).

### Naloxone Formulations

During the 210-month study period, three formulations of naloxone were predominantly distributed. For both vial sizes of the liquid injectable, following product labelling, participants were instructed that one dose is 1 mL administered intramuscularly. Intramuscular syringes (typically 25 gauge x 1” or 0.5mm x 25 mm) were provided in the kit; intravenous delivery of naloxone was almost never reported. Patient counselling and graphical printed cards advised that intramuscular administration could be achieved directly through clothing, into the shoulder or buttocks. These instructions have long been standardized among harm reduction programs nationwide, with many reproducing the same graphic developed at Chicago Recovery Alliance, and were consistent at Prevention Point Pittsburgh for the entire 17 years. In earlier years, one 10 mL vial could have been used in more than one overdose response event; by contrast, even with fractional dosing, 1 mL vials were not reported to be reused. For the nasal spray, one dose was defined as one full actualization in one nostril.

Naloxone provided in each kit:

- One 10 mL vial of 0.4 mg/mL naloxone hydrochloride: 2005 to 2015
- Two 1 mL vials of 0.4 mg/mL naloxone hydrochloride: October 2012 to January 2023
- Two units of 4 mg naloxone hydrochloride nasal spray: August 2016 to January 2023

Additional formulations available indirectly or briefly during the study period:

- 2 mL pre-filled needleless syringes of 1 mg/mL naloxone hydrochloride with aftermarket nasal adaptor [20]. This combination was not distributed by Prevention Point Pittsburgh, but some participants had received it elsewhere and reported using it when presenting for refills.
- 2 mg naloxone in 400 μL autoinjector. The autoinjector was briefly available in 2016 through a small donation of demonstration units from the manufacturer (Kaléo, Richmond, Virginia, US).

### Definitions

*Dose* was defined as the lowest single dose in approved labeling for overdose reversal. For 1 mL and 10 mL vials, a single dose was defined as 0.4 mg delivered intramuscularly, and 4 mg intranasally (in one nare) for the nasal spray.

*Overdose response events* (OREs) include any report of an attempted overdose reversal regardless of the outcome of the event (successful resuscitation, death, or unknown outcome) where naloxone was administered (or in one report attempted to be administered, see Death Case Review). While the term “reversals” is commonly used in the literature, “ORE” was considered a more accurate term in the context of these data.

*Cumulative utilization rates* are a quantification of how many units of dispensed naloxone were administered to reverse an overdose, during a specified calendar time period. They were calculated in two ways, both with the number of overdose response events as the numerator. The two denominators were either the number of units dispensed or standardized across formulations by the number of doses in a packaged unit. While this provides a useful metric for programmatic and financial planning, and as an input in modeling studies, it only accounts for doses dispensed by and OREs reported to Prevention Point Pittsburgh. Specifically, naloxone obtained via pharmacy or other harm reduction programs would not be accounted for in utilization rate denominators, and OREs not reported to the program would not be accounted for in the numerator.

“*Felt sick*” was understood by participants and staff to mean “dopesick” from precipitated opioid withdrawal. This is a different semantic meaning than generalized malaise recorded in spontaneous adverse event reporting systems.

*Titration* in the context of community distribution was the administration of fractional doses of liquid injectable naloxone, as reported by the respondent (e.g., “one and half doses”). Since only the total number of doses were recorded, there may have been instances of titration where two ½ doses were administered but would only appear as a single dose (1.0) in the data. Therefore, this metric should be considered to have high specificity, but with misclassification resulting in bias towards the null in adverse event analyses. Nasal spray and auto-injector formulations were not capable of delivering fractional doses in the manner dispensed.

### Statistical Analysis

In descriptive analyses, *t*-tests were used to assess between-group differences for continuous variables, with the Satterthwaite approximation for degrees of freedom [48] employed when unequal variance between groups was present, or Fisher’s exact test for low cell counts. Two-tailed chi-square distributions were used.

### Time-series Modeling

Data were aggregated by calendar month. Changes over time were assessed in four ways. First, smoothed monthly time trends (details in Data Visualization) were plotted along with 95% confidence intervals of the mean. Second, to summarize macro time differences between the beginning and end of the observation period, we calculated means within each level of categorical variables which compared the first 24 months to the last six months (Table 2). These timeframes were selected to balance the number of observations, as volume in later years was considerably greater than at the start. Third, record-level data were aggregated by calendar month for segmented regression, specifically piecewise linear regression, to identify abrupt changes in time trends empirically. The model optimized data fitting to a single breakpoint between two straight lines with differing slopes. It was implemented using ‘nl hockey’ in Stata MP version 17 (College Station, TX, USA). Results are summarized in Table 2 and detailed visualizations are in Supplemental Material. Fourth, based on programmatic context and policy implication discussions, further time-trend modeling used linear splines at pre-specified dates if a specific question about an expected changepoint was being evaluated, such as a law enacted on a certain date or a major change in service delivery. Splines were modeled using ‘mkspline’ and incorporated as mixed effects linear models using ‘xtmixed’ in Stata.

### Relative Dose

Relative dose was defined as a relative ratio of the number of doses of naloxone administered in an overdose response event, useful when comparing between formulations. First, average number of doses per ORE were calculated by formulation, reporting the arithmetic mean and 95% confidence interval of the mean. Consistently over 17 years, participants had been counselled that 1 mL equals one dose, regardless of whether it came from the 10 mL vial or 1 mL vial. Since the 1 mL vial had the lowest number of doses per ORE, it was selected as the reference group. Relative doses by formulation are reported as percent higher doses per ORE, and population averages calculated using scaled Poisson regression, assessed using two-tailed chi-square Wald tests.

### Rate Differences

To compare adverse event (AE) incidence rates (per 1,000 OREs), rate differences between formulations were also estimated using Poisson regression in Stata, adjusting for the number of doses as a linear continuous variable. Since the 1 mL vial had the lowest rates of adverse events, it was treated as the reference group; the rate difference represents the number of additional adverse events per 1,000 OREs that were observed with the 4 mg nasal spray or multiple formulations, relative to the number of AEs for the 1 mL vial. This metric provides a general measure of population level side effects for each of the major formulations; it is not meant to be interpreted as counterfactual inference as would result from a causal pharmacoepidemiology study where strength of conclusions could be drawn based on control for confounding by formulation.

### Data Visualization

In time series plots, calendar month is presented on the horizontal axis from July 2005 to January 2023 (n=211 months), except for adverse events in Figure 12 which are aggregated by year (2017-22) due to lower incidence and not having been collected in earlier years. The vertical axes are monthly arithmetic means. Overlaid scatterplots represent unadjusted data points. Time series were smoothed with locally estimated scatterplot smoothing (LOESS) and 95% confidence internal, with a 6-month window. In Supplemental Material for segmented regression, the two linear segments intervals are visualized instead. Because denominators for time trends can be different across figures, sparklines below the main time trends panel are provided to visualize fluctuations in monthly counts of new participants (Figures 2 and 5), naloxone doses distributed (Figure 6) or administered (Figure 10), or number of ORE (Figure 11). For visualizing the number of doses per ORE by formulation in Figure 9, a Gaussian (three sigma) smoother was applied. Figures were generated using Python 3.7 ‘matplotlib’ within a distributed Deepnote (deepnote.com) environment.

### Death Case Review

Prevention Point Pittsburgh conducted a review of reports in which the person administered naloxone was reported to have died. Paper records (encounter notes and written program logs) with interview notes were retrieved for the 23 deaths that were reported by participants who had been at the scene and who had administered naloxone (n=1,909) from 2020 to 2023; only one of these records did not contain any additional contextual information, but circumstances of death were present in the 22 others. Narratives of each death were constructed based on available records and assessed on when naloxone was administered relative to likely time of death, evidence of other causes of death described by health professionals, or other circumstances.

### Study Conduct

#### Open Science Practices

Pre-registration: DOI osf.io/b2f4h. Codebook, data collection form, and analytic code: DOI osf.io/sq5d6.

#### Institutional Ethics Review

This study was reviewed by the University of North Carolina-Chapel Hill Office of Human Research Ethics and deemed to be exempt, as anonymized secondary data research (22-2714).

#### Data Access

Data from Prevention Point Pittsburgh were provided to analysts on January 30, 2023. Data were anonymized so no individually identifiable information was available to analysts.

#### Participation of People with Lived Experience

People with lived experience of drug use, naloxone distribution, and overdose reversal were involved in the design and conduct of the services provided by Prevention Point Pittsburgh, as well as study conceptualization and interpretation of results. Results were to be reported back to participants of Prevention Point Pittsburgh through posters at program sites, a dedicated website, and a community presentation.

## RESULTS

### Overview

Descriptive quantitative results and time trend analysis are presented for each variable analyzed first. These are followed by programmatic context notes provided by Prevention Point Pittsburgh staff.

The first of five topic areas covers characteristics of participants receiving naloxone, which was deemed of high importance by harm reduction program co-authors. The subsequent four sections focus on biomedical and behavioral features: naloxone distribution, circumstances of overdose response events, response behaviors, and adverse events.

Trends in naloxone distribution and administration were examined from July 24, 2005 through January 24, 2023, comprising an observation period of 211 consecutive calendar months (inclusive), or 17.6 years. A total of 16,904 service encounters by 7,582 unique participants, resulted in 70,234 doses of naloxone dispensed, with 5,521 OREs reported, utilizing 8,756 doses of naloxone.

### New Participant Volume

#### Quantitative Results

After a decade of steady volume (Figure 1), unique new participant intakes increased sharply starting January 2015. Pennsylvania Act 139 allowed standing orders and third-party naloxone prescriptions, and Prevention Point Pittsburgh’s Medical Director issued a standing order for the organization, enabling naloxone distribution without requiring individual prescriptions. Monthly new participant intakes increased from an average of 10.4 per month (95% CI: 9.4, 11.4) to 65.9 per month (95% CI: 60.7, 71.1) after the law was enacted, translating to 56.2 (95% CI: 46.5, 65.8; Wald *χ*^2^ 510, 3 df, p<0.001) additional new participants per month (Figure S1).

**Figure 1.**
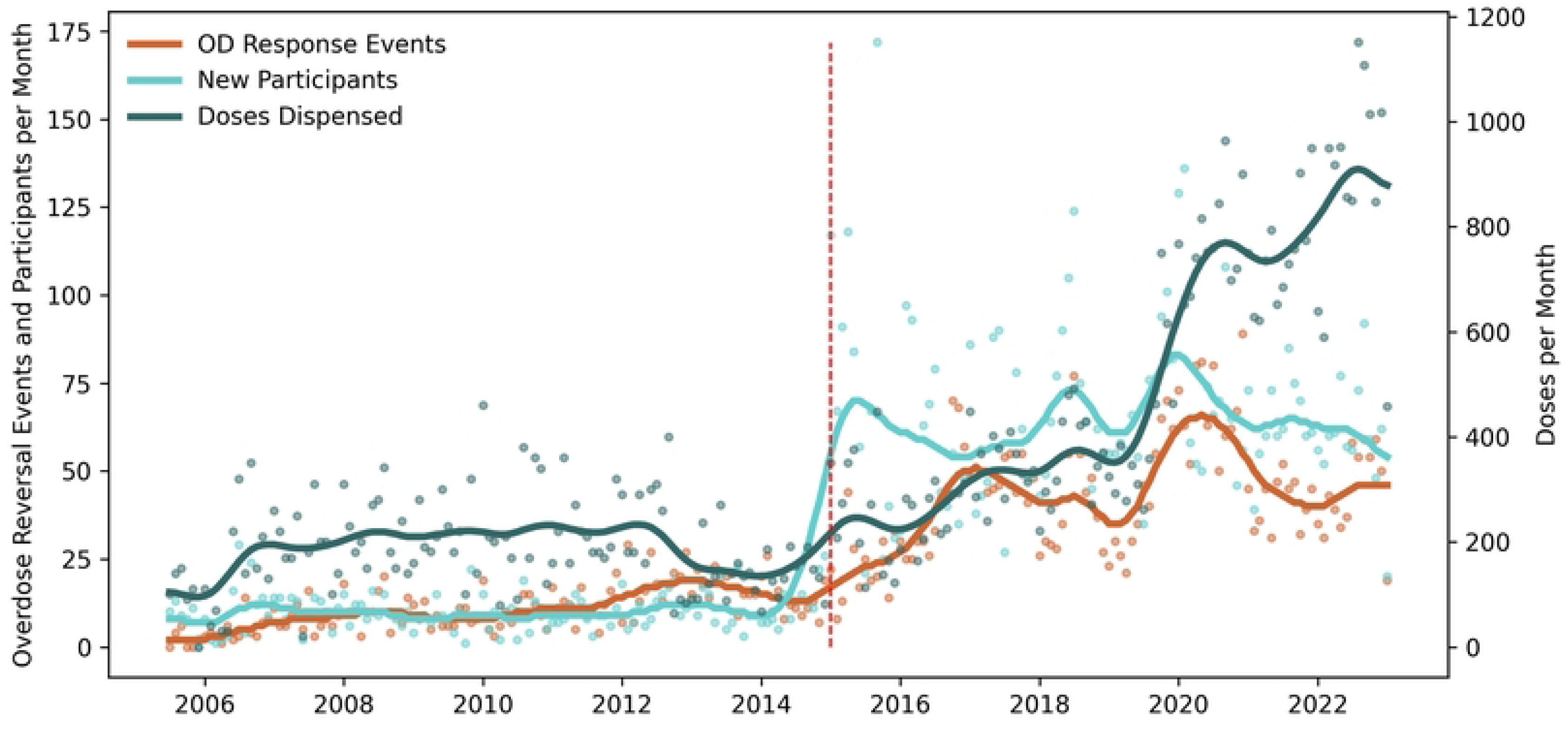
Monthly volume of new participants trained, naloxone dispensed, and overdose response events reported. Dots are raw monthly counts; lines are moving averages. Three time series are presented on two vertical axes. The left vertical axis is the number of new participants trained and the number of monthly overdose response events reported. The right vertical axis is the monthly count of doses of naloxone dispensed. The vertical red dashed line at December 2014 represents the change in state legislation.

However, despite the immediate increase in new participants, the utilization of naloxone was lower. Average ORE per new participant enrolled in 2014 was 1.46 (95% CI: 0.84, 2.1), but in 2015 after the law change, it fell to 0.47 (95% CI: 0.24, 0.70). This decrease mirrors, in part, a downward secular trend in OREs from 2013-2015 following the emergence of illicitly manufactured fentanyl in 2013, Figure 2. However, though a similar peak in overall OREs occurred in 2016-2017 as carfentanil was known to be circulating, no concurrent spike in new participant OREs occurred after enactment of state legislation. Overall, the trendline in OREs reported by new participants following the law change no longer mirrored the overall trendline, a decoupling that may suggest that new participants tended to be qualitatively different before and after the legislation.

**Figure 2.**
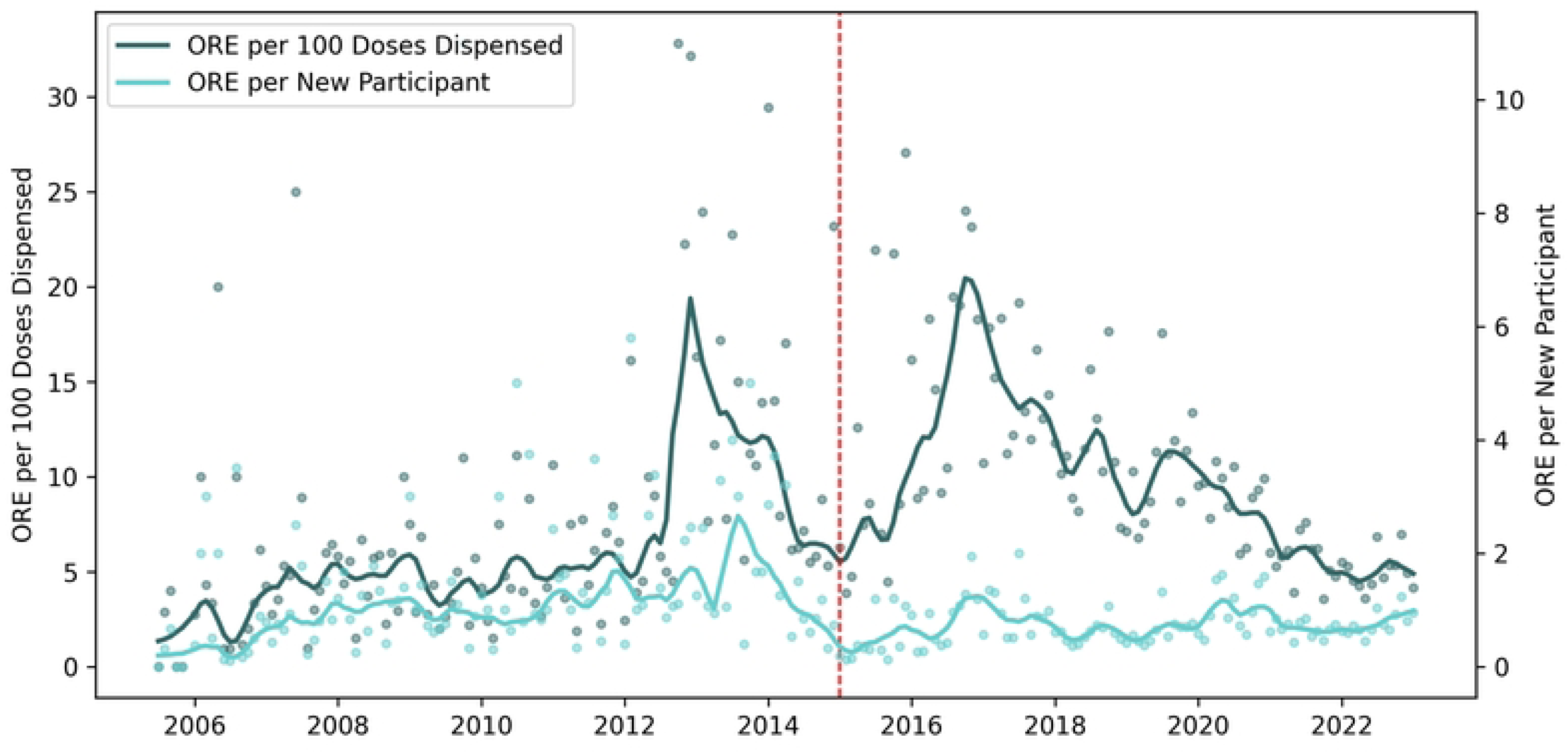
Monthly Rates of Overdose Response Events per 100 Doses of Naloxone Dispensed and New Participants. Dots are monthly rates; lines are moving averages. Monthly utilization rates of naloxone during overdose response events (ORE) are presented using doses dispensed and new participants as denominators. Red line at December 2014 represents the enactment of legislation enabling community based naloxone distribution. Study dates: July 2005 to January 2023.

#### Programmatic Context

Program staff verified the quantitative finding that the volume of new participants increased immediately after the law was enacted. However, they also provided context that, compared to previous years, new participants were demographically different: Increases were disproportionately in patients more likely to be older, female and of White race. In early 2015, the venues for overdose prevention trainings shifted from syringe services sites to fire houses and community centers, where audiences were exclusively constituted of the concerned general public, and not people who used drugs themselves. Program staff reported initial excitement at the broader reach of the intervention, but a few months later began to realize that the newly trained general community members were not reporting naloxone utilization or reversals.

Accordingly, program staff felt that demographic changes after enactment of the law were crucial to document because they have direct implication on public health practice. Therefore, we proceeded to quantify the programmatic observation that the law led to a change in the underlying population receiving naloxone at Prevention Point Pittsburgh.

### Participants: Age

#### Quantitative Results

The median age at the initial training was 40 years (IQR: 31 years, 53 years), Table 1. Segmented regression (Table 2, Figure S3) identified a breakpoint in April 2015. From 2005 to 2015, average age increased in linear single-year increments during the first decade of naloxone distribution (Figure 3), so that the average age in 2005 was 31.6 years, and 41.5 in 2015, suggesting a possible birth-cohort influence if the source population is assumed to be stable without replenishment. Immediately following the enactment of the legislation, new participants were older: The average age increased from a baseline 37.0 years-old (95% CI: 36.3, 37.7) to 46.0 years-old (95% CI: 45.0, 46.9, *t-*test 14.9, 2048 df, *p*<0.001) the following year.

**Figure 3.**
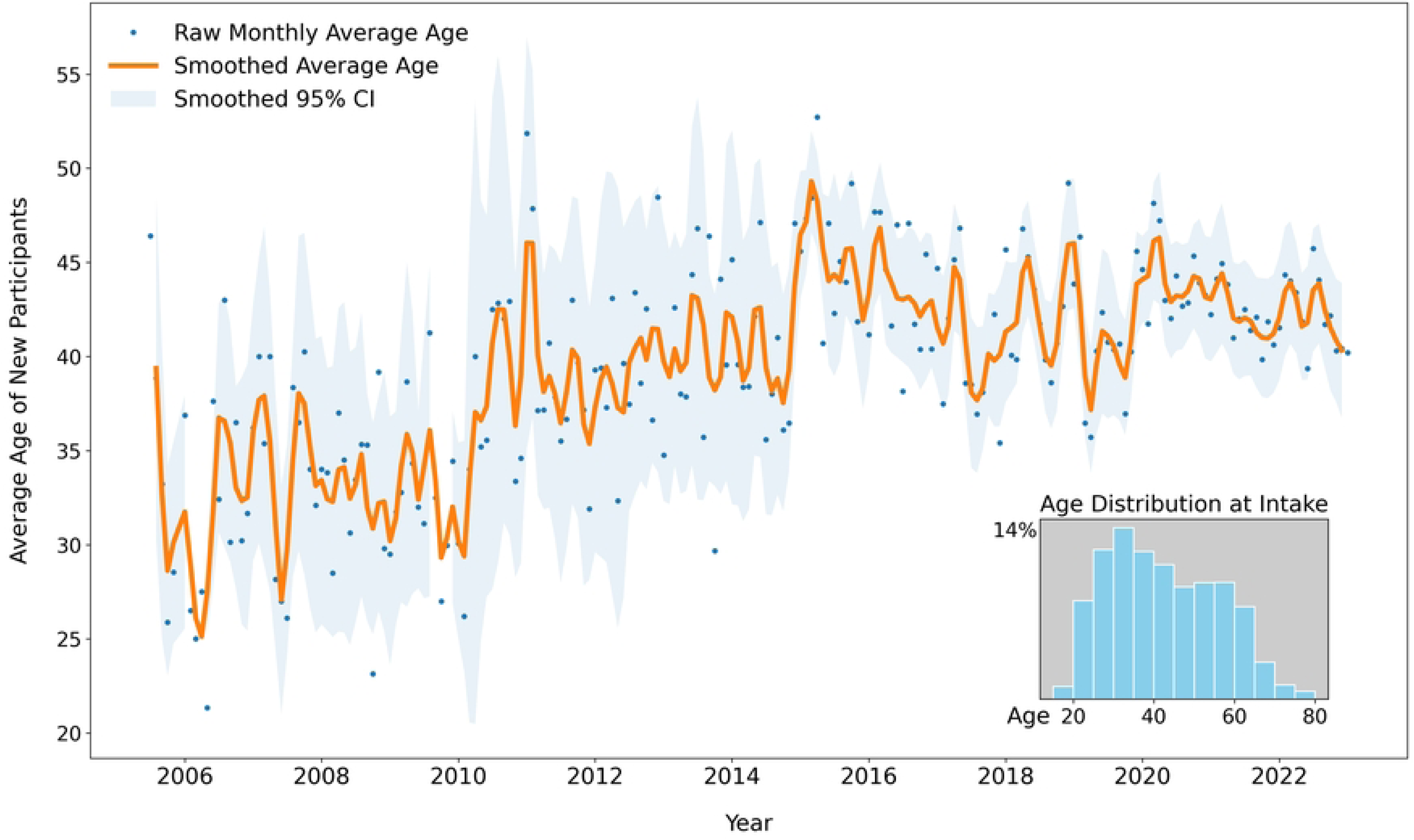
Average age of new participants receiving overdose reversal training and naloxone. Dots represent average age of new participants, by month. Time series of monthly average age for new participants receiving initial overdose prevention training and naloxone dispensing. Time series line (orange) and shaded 95% confidence interval of the mean (light blue) have been smoothed, and raw average age per month is depicted as dots. Inset plot is a histogram of average age of new participants, by 5-year bin increment, during the entire observation period: July 2005 to January 2023.

**Table 1.**
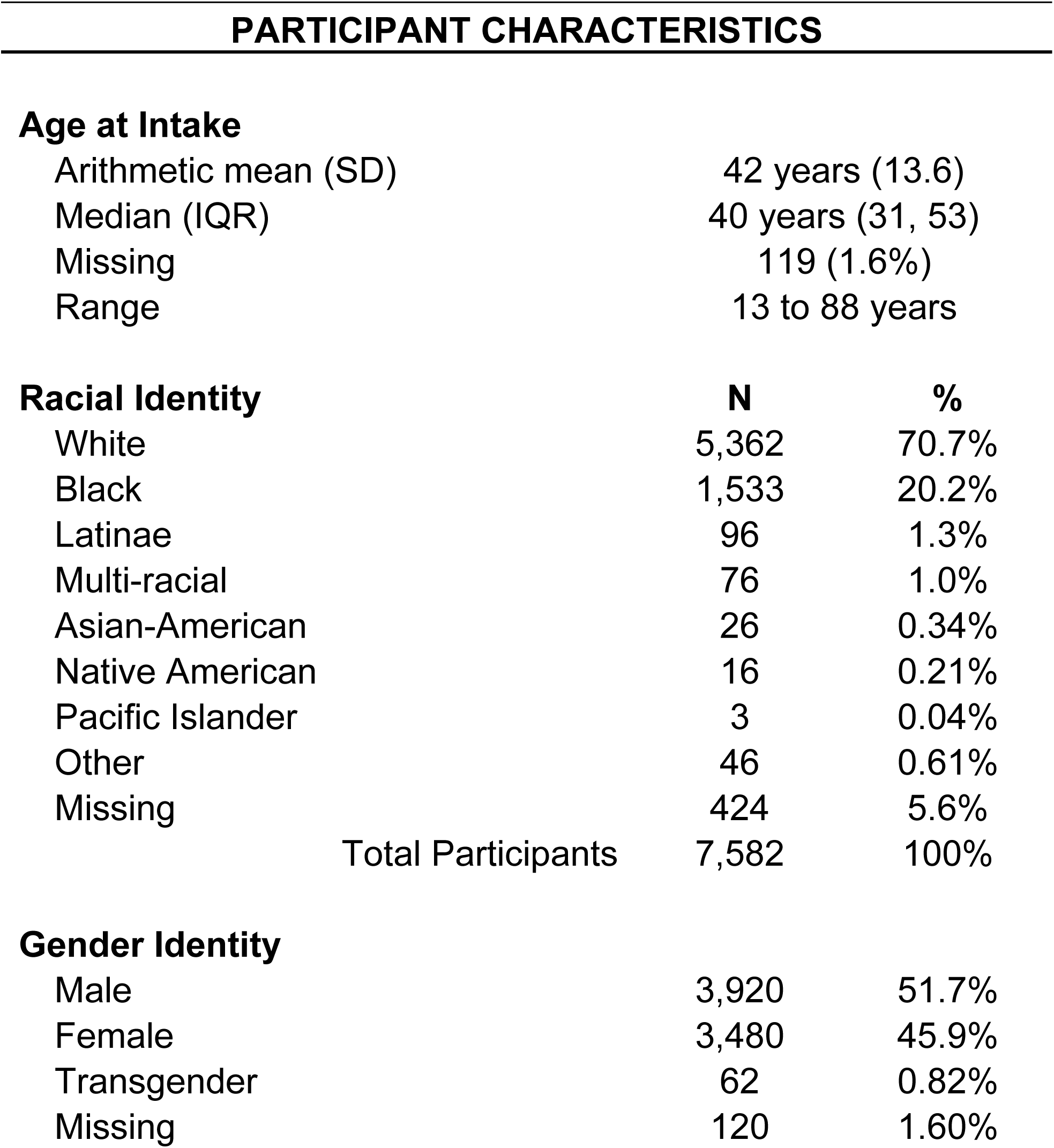
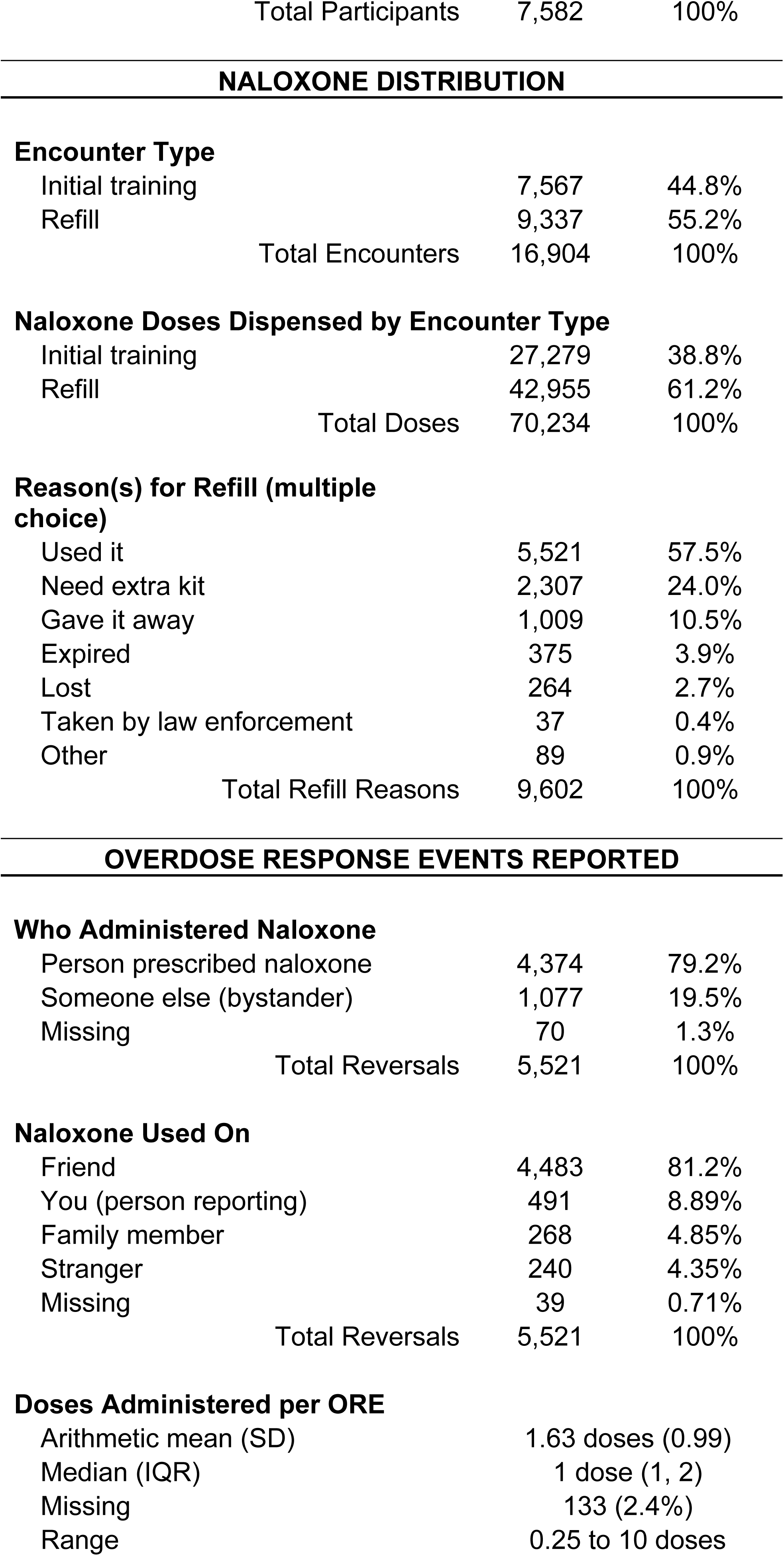
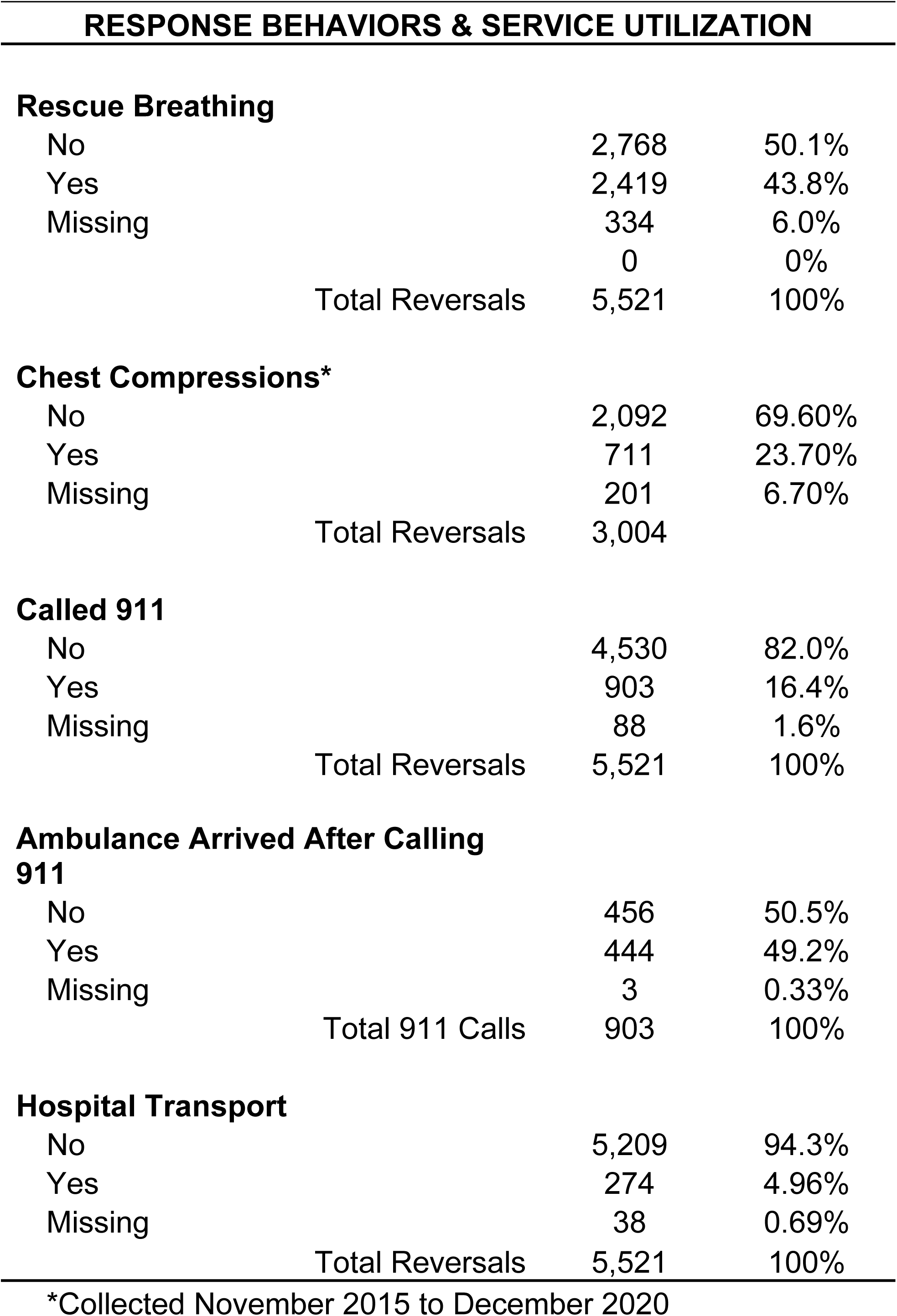
Participant Characteristics, Behaviors, and Service.

| PARTICIPANT CHARACTERISTICS |  |  |
| --- | --- | --- |
| **Age at Intake** |  |  |
| Arithmetic mean (SD) | 42 years (13.6) |  |
| Median (IQR) | 40 years (31, 53) |  |
| Missing | 119 (1.6%) |  |
| Range | 13 to 88 years |  |
| **Racial Identity** | **N** | **%** |
| White | 5,362 | 70.7% |
| Black | 1,533 | 20.2% |
| Latinae | 96 | 1.3% |
| Multi-racial | 76 | 1.0% |
| Asian-American | 26 | 0.34% |
| Native American | 16 | 0.21% |
| Pacific Islander | 3 | 0.04% |
| Other | 46 | 0.61% |
| Missing | 424 | 5.6% |
| Total Participants | 7,582 | 100% |
| **Gender Identity** |  |  |
| Male | 3,920 | 51.7% |
| Female | 3,480 | 45.9% |
| Transgender | 62 | 0.82% |
| Missing | 120 | 1.60% |
| Total Participants | 7,582 | 100% |
| Encounter Type |  |  |
| Initial training | 7,567 | 44.8% |
| Refill | 9,337 | 55.2% |
| Total Encounters | 16,904 | 100% |
| Naloxone Doses Dispensed by Encounter Type |  |  |
| Initial training | 27,279 | 38.8% |
| Refill | 42,955 | 61.2% |
| Total Doses | 70,234 | 100% |
| Reason(s) for Refill (multiple choice) |  |  |
| Used it | 5,521 | 57.5% |
| Need extra kit | 2,307 | 24.0% |
| Gave it away | 1,009 | 10.5% |
| Expired | 375 | 3.9% |
| Lost | 264 | 2.7% |
| Taken by law enforcement | 37 | 0.4% |
| Other | 89 | 0.9% |
| Total Refill Reasons | 9,602 | 100% |
| Who Administered Naloxone |  |  |
| Person prescribed naloxone | 4,374 | 79.2% |
| Someone else (bystander) | 1,077 | 19.5% |
| Missing | 70 | 1.3% |
| Total Reversals | 5,521 | 100% |
| Naloxone Used On |  |  |
| Friend | 4,483 | 81.2% |
| You (person reporting) | 491 | 8.89% |
| Family member | 268 | 4.85% |
| Stranger | 240 | 4.35% |
| Missing | 39 | 0.71% |
| Total Reversals | 5,521 | 100% |

Doses Administered per ORE
|  |  |
| --- | --- |
| Arithmetic mean (SD) | 1.63 doses (0.99) |
| Median (IQR) | 1 dose (1, 2) |
| Missing | 133 (2.4%) |
| Range | 0.25 to 10 doses |
| **Rescue Breathing** |  |  |
| No | 2,768 | 50.1% |
| Yes | 2,419 | 43.8% |
| Missing | 334 | 6.0% |
|  | 0 | 0% |
| Total Reversals | 5,521 | 100% |
| **Chest Compressions\*** |  |  |
| No | 2,092 | 69.60% |
| Yes | 711 | 23.70% |
| Missing | 201 | 6.70% |
| Total Reversals | 3,004 |  |
| **Called 911** |  |  |
| No | 4,530 | 82.0% |
| Yes | 903 | 16.4% |
| Missing | 88 | 1.6% |
| Total Reversals | 5,521 | 100% |
| **Ambulance Arrived After Calling 911** |  |  |
| No | 456 | 50.5% |
| Yes | 444 | 49.2% |
| Missing | 3 | 0.33% |
| Total 911 Calls | 903 | 100% |
| **Hospital Transport** |  |  |
| No | 5,209 | 94.3% |
| Yes | 274 | 4.96% |
| Missing | 38 | 0.69% |
| Total Reversals | 5,521 | 100% |

\*Collected November 2015 to December 2020

**Table 2.**
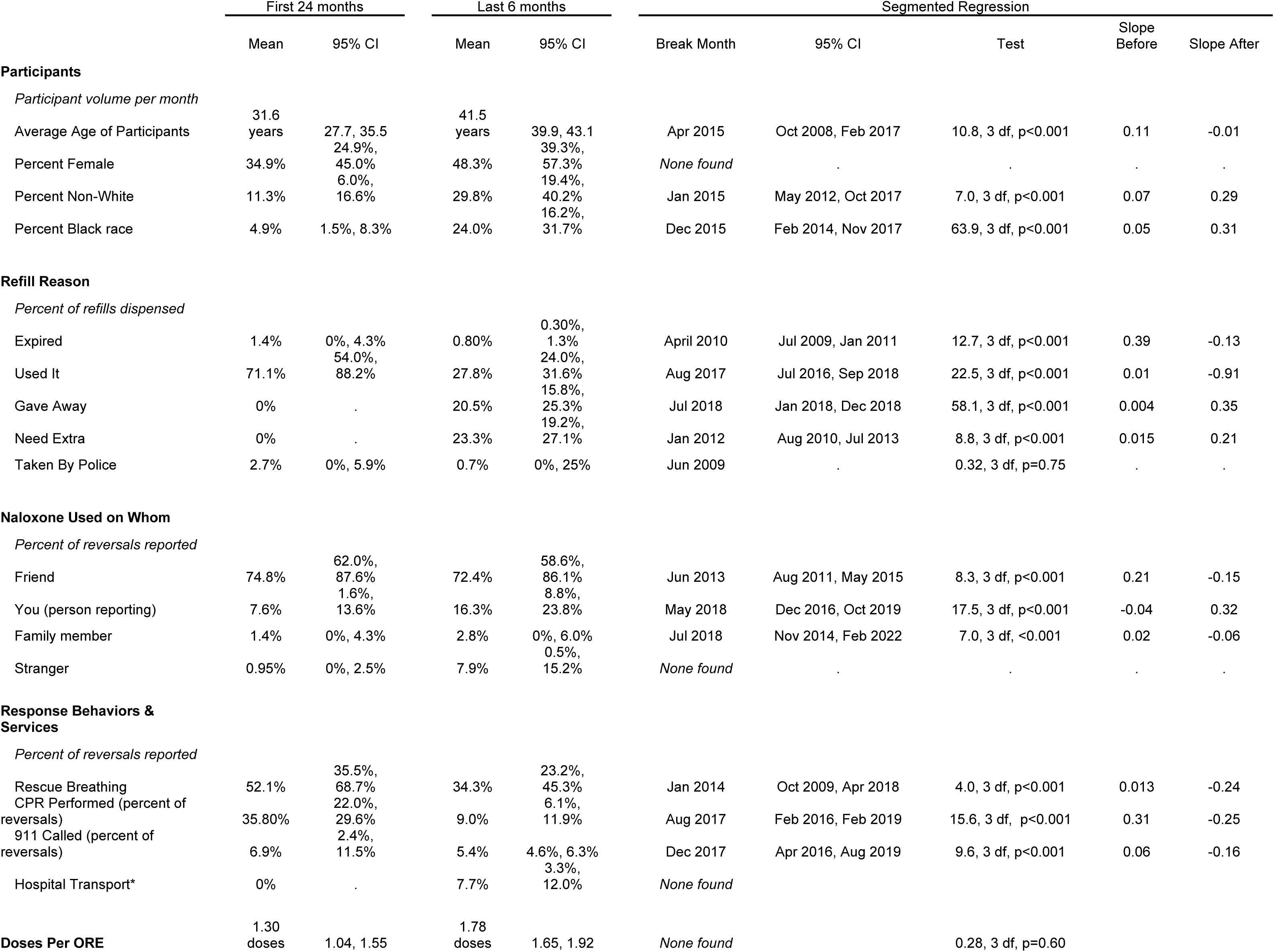
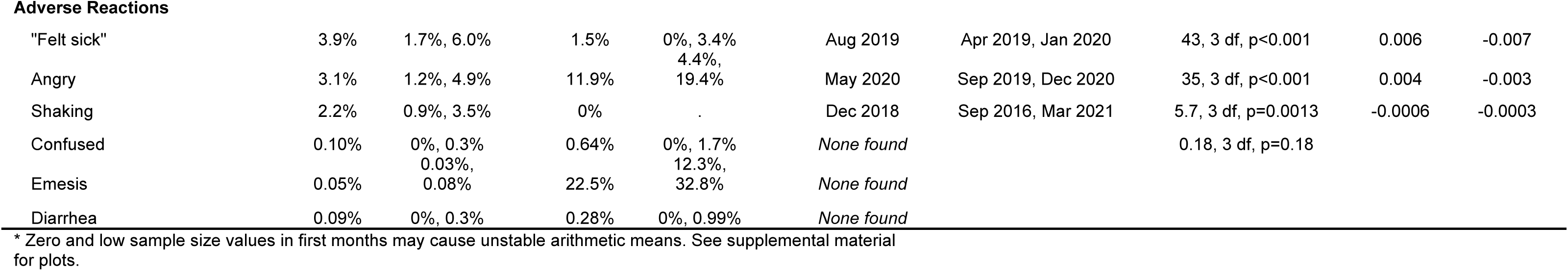
Participant characteristics, behaviors, and adverse reactions in first 24 month and last 6 months, with segmented regression breakpoints Participants.

#### Programmatic Context

Program staff observations corroborated quantitative findings. The immediate increase in age during 2015 was due to large-audience naloxone training events hosted by Prevention Point Pittsburgh that attracted parents concerned about their children. These events were often held in venues outside of Pittsburgh city limits [49], including suburban and rural areas of western Pennsylvania. In a departure from traditional service delivery, these large-audience events attracted more family and friends than people who use drugs.

This increase in terms of age leveled out by the end of 2016 as Prevention Point Pittsburgh refocused outreach efforts to serve people who use drugs instead of concerned friends and family. This change in focus was made intentionally by program staff in early 2016 because there were fewer-than-expected ORE reports despite the large increase in new participants. Program staff believed that naloxone distributed in the mass training events to friends and family were less likely to be used, compared to when distributed directly to people who use drugs. In the most recent data years (2022-3), the downward age trend was explained as being due to programmatic expansion to mobile sites in previously underserved neighborhoods drawing younger populations.

### Participants: Gender Identity

#### Quantitative Results

Self-reported gender identity was 51.7% male, 45.9% female, and 0.82% (n=62) transgender during the entire observation period. Although segmented regression did not identify a single breakpoint, visual inspection of the timeline (Figure 4) and discussion with Prevention Point Pittsburgh staff suggested that the percent of new participants identifying as female increased in 2015-16, immediately following the enactment of the aforementioned naloxone law. The percent of female new participants increased from 34.6% (95% CI: 31.4%, 37.8%) to 58.1% (52.7%, 63.5%, *t-*test 8.0, 22 Satterthwaite df, *p*<0.001) in the calendar year before and after legislation.

**Figure 4.**
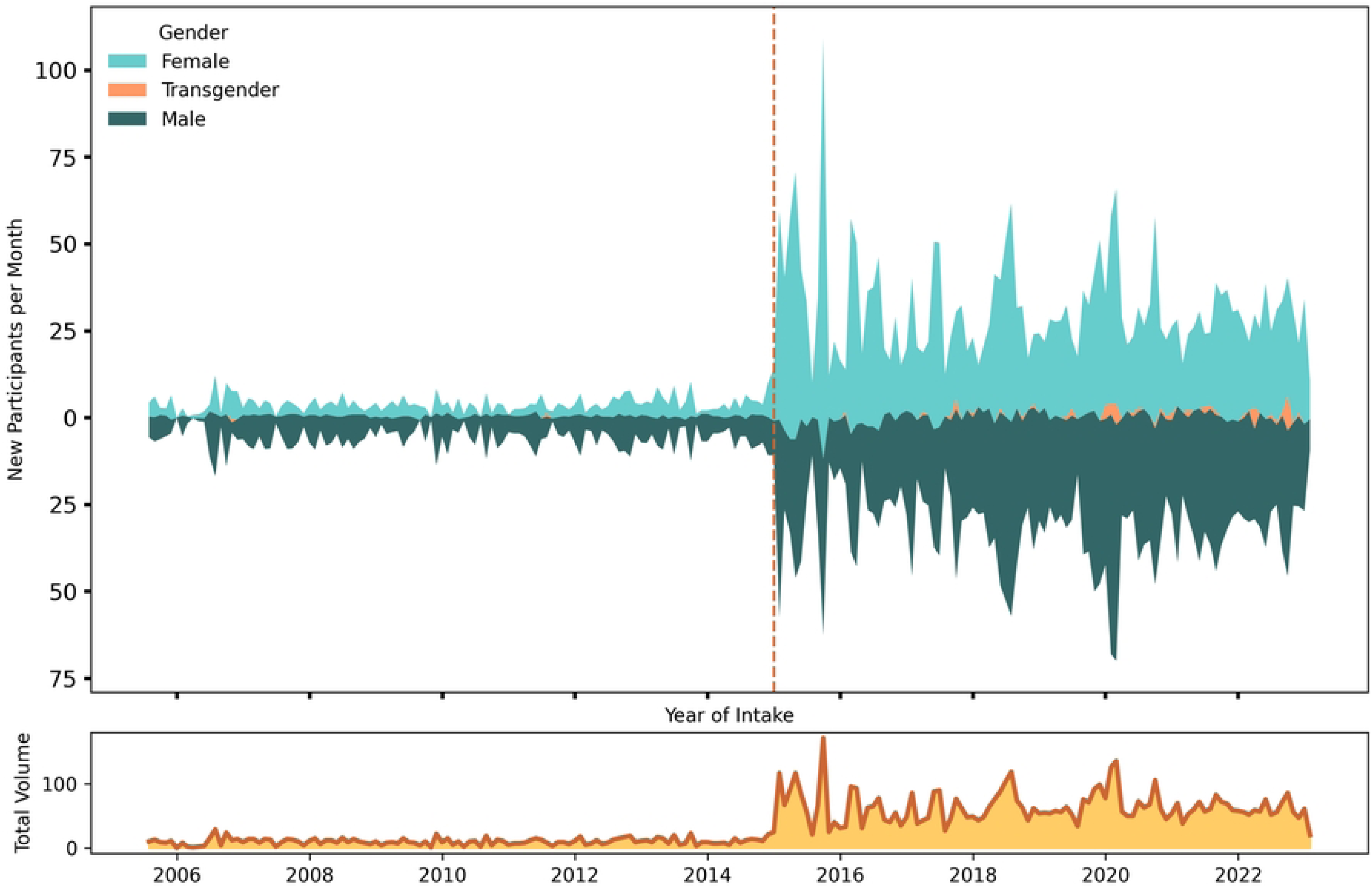
New participant volume and self-reported gender for naloxone training and dispensing. Top frame depicts timeline in new participant volume receiving naloxone at initial training encounters, by self-reported gender. River plots are interpreted as stacked area graphs, with area above and below zero both being in the “positive” direction by category. The vertical red dashed line at December 2014-to-January 2015 interface represents the change in state legislation. Bottom frame is a smoothed sparkline of total volume of all new monthly participants. Study dates: July 2005 to January 2023.

#### Programmatic Context

Quantification confirms observations by Prevention Point Pittsburgh staff that after the legislation there was increased representation of concerned females (often mothers of children who use opioids) who attended large training events. After 2016, Prevention Point Pittsburgh made the conscious decision to re-prioritize people who are actively using drugs for naloxone outreach, and the percent female new participants stabilized to around 40-50% per month. In recent years (2022-23), there was a noticeable uptick in the proportion of females to 50-60%, concurrent with the younger average age noted previously. This period is contemporaneous with a shift to nasal naloxone formulations distributed and expansion of mobile sites to new Pittsburgh neighborhoods, with the caveat that this association may be incidental.

### Participants: Racialized Identity

#### Quantitative Results

Self-reported racialized identity of participants was 70.7% White, 20.2% Black, 1.3% Latine, 1% multiracial, with less than 1% each of others, Table 1. The racial distribution mirrored the population of Allegheny County, with greater representation by individuals identifying as Black, and fewer of Latine origin [50]. Unlike age and gender, the impact of the state legislation on race was less immediately evident and more nuanced. Segmented regression identified an inflection point in January 2015, showing an increase in non-White new participants (Table 2, Figure S4). However, segmented regression revealed that the increase in Black new participants did not actually start until a year later, after the end of December 2015 (Table 2, Figure 5).

**Figure 5.**
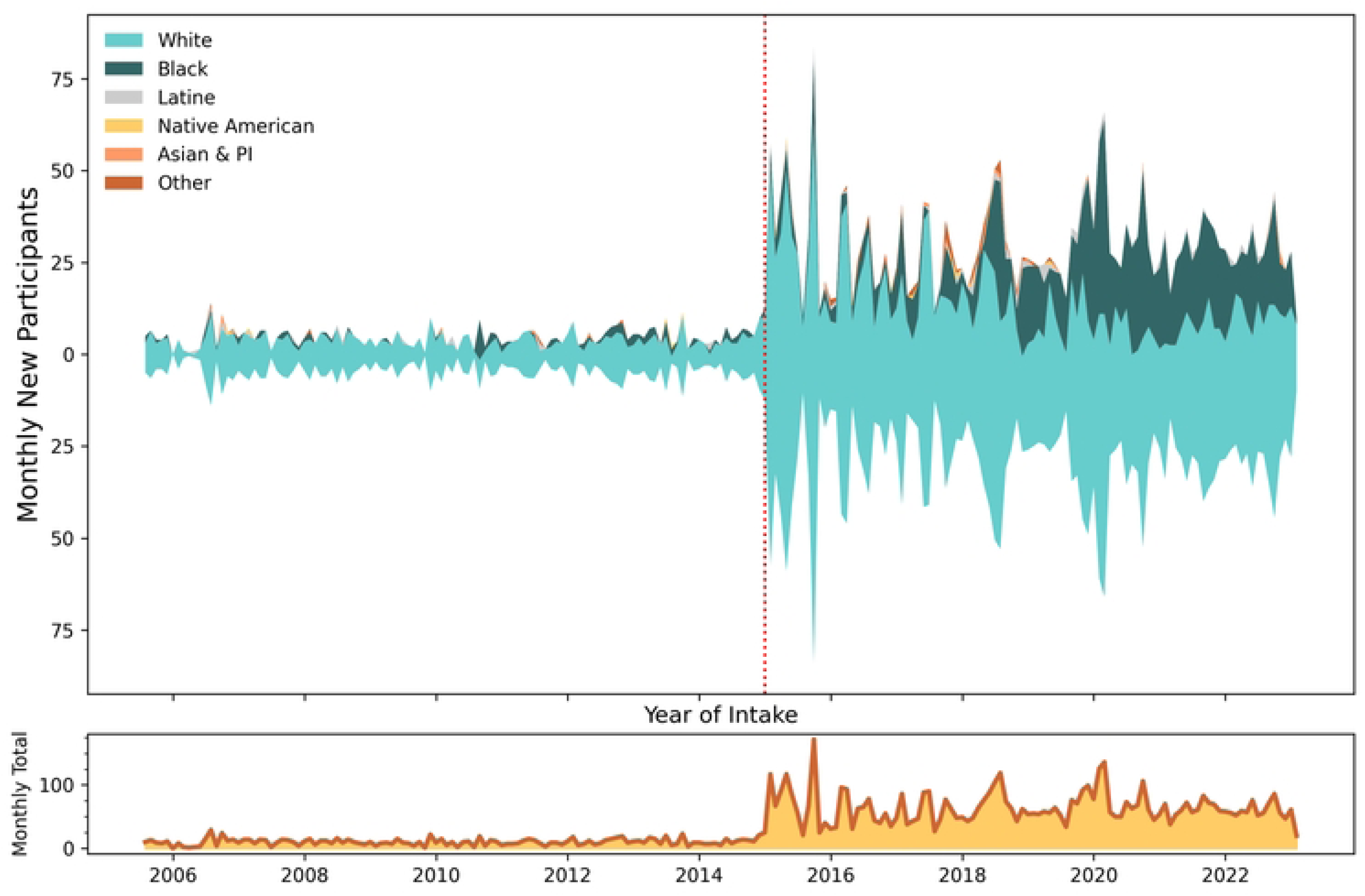
Racial identity of new participants dispensed naloxone. Top frame depicts timeline in new participant volume to naloxone training and dispensing, by self-reported racial identity. The vertical red dashed line at December 2014 represents the change in state legislation. Bottom frame is a smoothed sparkline of total volume of all new monthly participants. Study dates: July 2005 to January 2023.

#### Programmatic Context

Prevention Point Pittsburgh staff noted that the increase in Black participants after 2015 was not surprising and was planned. A retrospective analysis of the first ten years of service delivery was conducted by staff in 2016 [16], which starkly quantified what up until then had been a casual observation, namely that the predominant race of participants was White, and that different strategies would be needed to engage people of color.

The statistical breakpoint in January 2016 conforms with a significant service change: In February 2016, Prevention Point Pittsburgh intentionally increased outreach to a predominantly Black neighborhood via mobile-based services and hiring community health advocates from the community to do naloxone distribution. A second predominantly Black neighborhood was served starting in late September 2019. While the law itself did not lead to a passive increase in Black participants, Prevention Point Pittsburgh staff did credit the law in allowing them to distribute naloxone in more places. For example, a barber shop in a predominantly Black neighborhood served as an early expansion site for naloxone distribution, a partnership that was brokered with the support of a local community leader identified by program staff. Staff further noted this active outreach was different than inbound solicitations from community groups for naloxone trainings; people who sought out Prevention Point Pittsburgh for group trainings tended to be serving White communities.

#### Iterative Modeling and Further Contextualization

Based on explanations by Program staff, time trend analysis was refined. Spline models with knots at the dates of law enactment and the two outreach expansion dates demonstrated good fit for these inflection points; however, the period from September 2019 to January 2023 displayed non-linear temporal distribution (Figure S5). Visual inspection was corroborated empirically by the predicted output from the mixed effects model, which showed that the percent of Black new participants increased considerably after September 2019 (ŷ_September_ = 27.0% to ŷ_October_ = 39.2%).

However, time-restricted segmented regression (Figure S4) identified a potential change point of December 2020 after which a decline in the percent of Black participants accelerated through the end of observation in January 2023, a result of expanded mobile services in another Pittsburgh neighborhood with a younger, predominantly White population. This particular mobile site was characterized by physicians who could start low-threshold buprenorphine inductions (i.e., a flexible treatment approach with same-day initiation, relaxed adherence requirements, and availability in non-traditional settings) [51] in the field. Low-threshold buprenorphine provision also drew participants to the harm reduction service side of the program, where take-home naloxone was also provided. Therefore, the increase in younger, White participants was the combined function of local demographics and the spillover effect stemming from providing medications for opioid use disorder.

In summary, enactment of a law in Pennsylvania led to an immediate increase in naloxone dispensed to concerned family and friends of people who use drugs. The law enabled Prevention Point Pittsburgh to expand to underserved neighborhoods, but inclusion of more racialized minorities also required service delivery innovation.

### Naloxone Distribution

#### Quantitative Results

During the 17-year period, half of all naloxone doses dispensed by Prevention Point Pittsburgh were in 1 mL (0.4 mg/mL) vials (n=35,715; 50.8%), followed by 10 mL (0.4 mg/mL) vials (n=18,420; 26.2%), and 4 mg nasal spray (n=16,063; 22.9%). Only 36 doses of the autoinjector were dispensed during a one-month period in 2016 with donated product. During the first decade of operation (Figure 6), Prevention Point Pittsburgh distributed the 10 mL vial exclusively.

**Figure 6.**
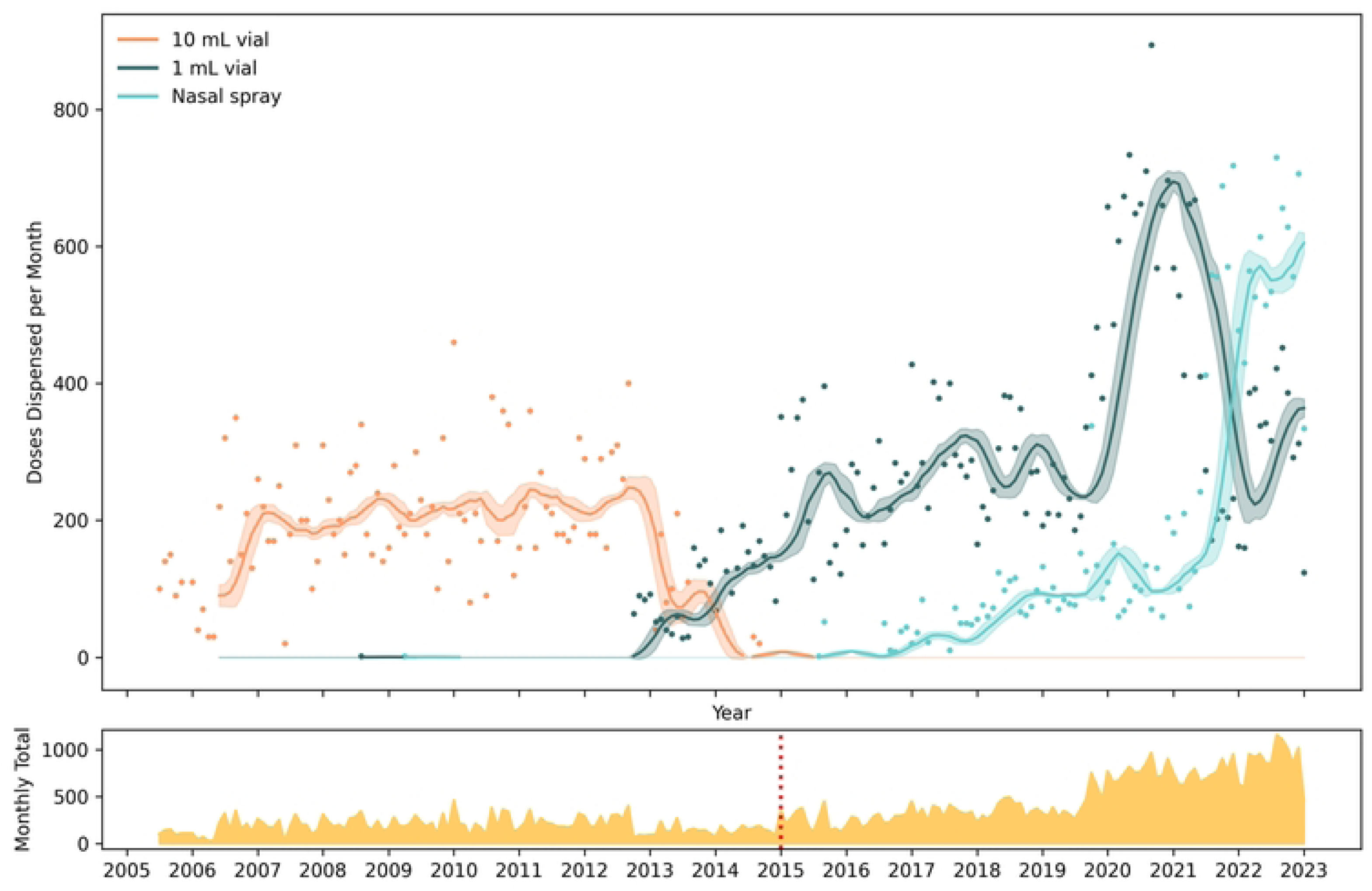
Major forms of naloxone distributed, July 2005 to January 2023. In the top panel, smoothed time series are displayed along with 95% confidence intervals of doses dispensed per month, by formulation. Colored dots represent raw counts of doses by month. The bottom panel is a sparkline showing total number of doses per month of naloxone distributed across all formulations. Initially only 10 mL vials were available for distribution. The vertical red dashed line at December 2014 represents the change in state legislation. The only other form of naloxone distributed by Prevention Point Pittsburgh was 36 units of an auto-injector in June 2016, which are not depicted. Study dates: July 2005 to January 2023.

#### Programmatic Context

Program staff provided context for the two points where lines crossed. Initially the 10 mL vial was the only formulation available, but the 10 mL vial was replaced by 1 mL vials starting in October 2012 due to a new contract with the manufacturer. With this transition, by August 2013 all IM distribution was of 1 mL vials, packaged by the program as kits containing two 1 mL vials. The other line crossing occurred in Spring 2021 when a manufacturing problem disrupted 1 mL vial production. From May 2021 to September 2022, a manufacturing shortage of affordable naloxone led to a shortage of the 1 mL vials. The State of Pennsylvania was able to increase bulk nasal naloxone to Prevention Point, but the organization had to hire a part-time staff person to help other community-based programs with accessing the state ordering portal. After the shortage was resolved, 1 mL vial purchases resumed in late 2022.

### Reasons for Refill

#### Quantitative Results

Over the 17.6 years, there were 16,904 participant-encounters by Prevention Point Pittsburgh staff, of which 44.8% (n=7,567) were initial trainings (Table 1). The remainder of dispensing events occurred when refill requests participants who had previously been trained returned for a refill.

A total of 70,234 doses of naloxone were dispensed: 27,279 during initial training encounters (38.8%) and 42,955 (61.2%) during refills. The average number of doses dispensed per participant was 4.15, cumulatively.

“Used it” was the most common reason for refill in 57.5% of refill requests (Table 1, Figure 7). Segmented regression revealed that prior to August 2017, about 70% of refill requests were after naloxone had been used (Table 2, Figure S8). After this date, participants increasingly asked for refills for other reasons.

**Figure 7.**
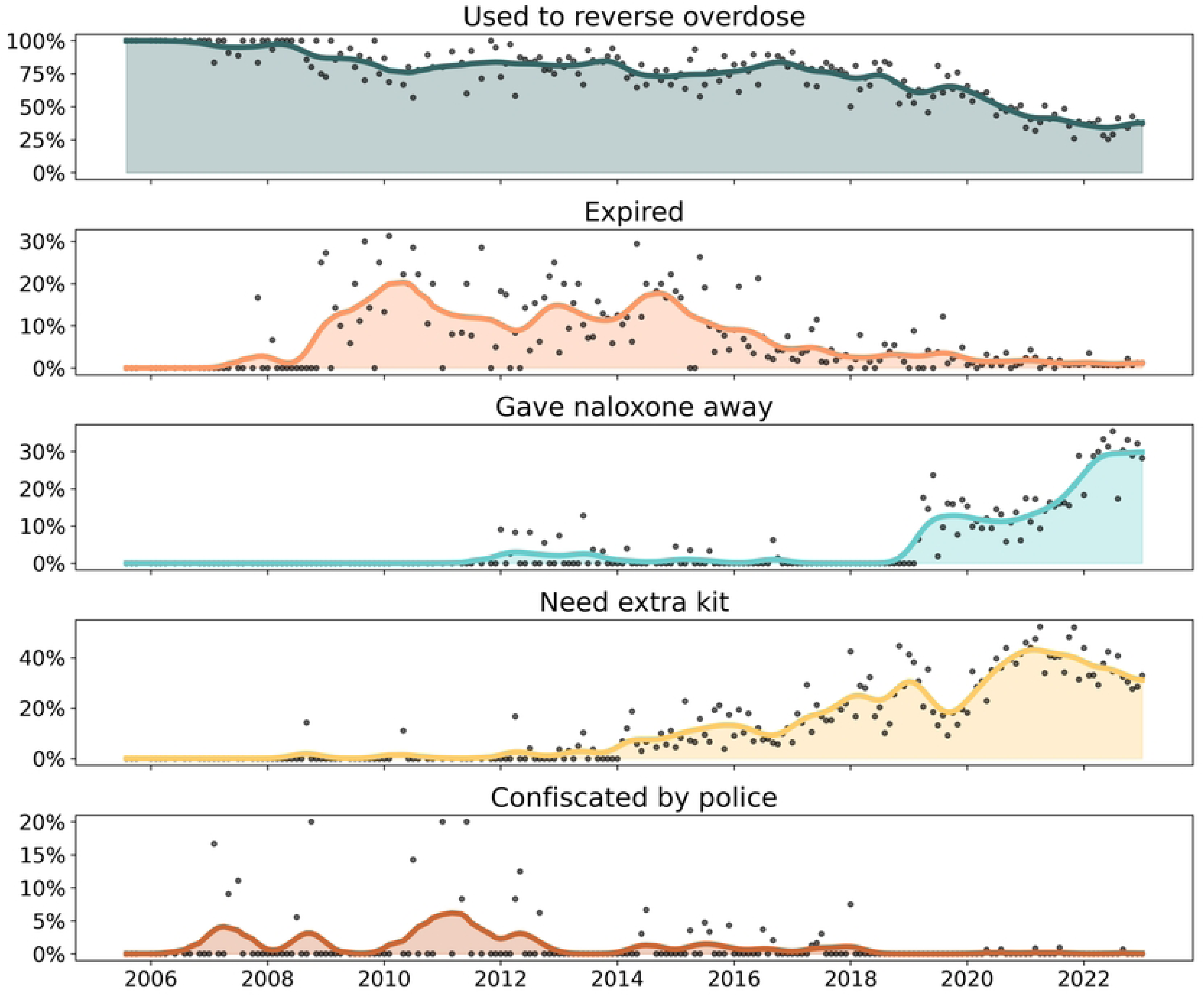
Reasons for requesting naloxone refills. Smoothed sparklines depicting monthly proportion of refill requests by reason for refill. Grey dots represent raw counts by month, while the colored lines are smoothed monthly average. Study dates: July 2005 to January 2023.

Obtaining a refill for naloxone because of expiration date was relatively rare, at 3.9% of refill requests overall. Segmented regression found an initial decrease in expired naloxone after April 2010 (Table 2), but inspection of trendlines (Figure 7) reveal further peaks over time, finally stabilizing in a lower proportion of refills from 2017 or 2018 onwards. Comparing these findings to Figure 6, the likelihood of expired naloxone refill does not correspond to the switch from 10 mL vials to 1 mL vials in calendar years 2012 to 2013. The further decline in proportion of expired naloxone in later years (2017 onwards) is somewhat contemporaneous with availability of the nasal spray, but samples are too small to draw further conclusions.

“Need an extra kit” was used by program staff to record when individuals who already had a kit and wanted a second one. This comprised 24.0% of refill requests overall, and segmented regression identified January 2012 as a change point (Table 2, Figure S7). Cumulatively, about 30% of refill requests were for this reason. This does not include people who used a kit and were looking for a refill.

“Gave away” increased substantially starting around July 2018 (Table 2, Figure S6), as suggested by segmented regression. While this reason was only 10.5% cumulatively, nearly a quarter of all refill requests were because of secondary distribution (“gave away”). Secondary distribution is a well-documented phenomenon where participants directly accessing harm reduction services obtain additional supplies for distribution among their peer networks informally [52,53].

Law enforcement confiscation of naloxone occurred mostly before the enactment of the statewide standing order in December 2014, in 37 instances. Other reasons for refill were very rarely reported (less than 1% combined), including only one or two reports of breakage of glass vials.

#### Programmatic Context

Program staff clarified that, following the law change, increased participant reports of sharing naloxone with others (“gave away”) may not represent a true change in behavior, but rather a gradual increase in participants’ level of comfort reporting giving away naloxone as a form of secondary distribution, a phenomenon documented elsewhere after law changes [54]. Prior to the law change and the official sanction of naloxone distribution, coded language was often used by participants to emphasize the *personal* need for the naloxone because third-party prescribing was not legally sanctioned. Program staff attributed the legacy of prescription status and the requirement to have a documented clinical encounter as having conditioned participants to only talk about needing naloxone for themselves. As third-party prescribing of naloxone became less legally risky over time, participants became more willing to candidly discuss that they were giving their naloxone kits away to others. Prevention Point Pittsburgh staff noted a de-stigmatizing of naloxone after the passage of the law, an advance that was critical to enabling more honest dialogue with participants about their experiences.

Program staff provided more context for “need an extra kit” responses. This category often refers to individuals wanting to have naloxone available in multiple locations, such as both at home and in cars, or one for home and one to carry in a handbag.

### Naloxone Administration

Naloxone administration reports were limited to those instances where naloxone had been used (n=5,521), instead of refill requests for other reasons. Naloxone was generally *administered* by the person to whom it had been prescribed and trained in 79.2% (n=4,374) of OREs, leaving 19.5% on average administered by “someone else” at the scene (Table 1). However, this was different (Wald *χ*^2^=218, df 5, p<0.001) by formulation: 33.2% of OREs with nasal sprays were done by someone else, whereas intramuscular formulations ranged from 10.1% to 16.4%. This suggested the necessity to examine to whom the naloxone was administered.

### Naloxone Used on Whom

#### Quantitative Results

The largest category represents 81.2% of ORE that were performed on friends and acquaintances, which decreased slightly over time (Figure 8, Table 1). Segmented regression yielded June 2013 as a change point after which other categories of people slightly increased in being administered naloxone instead of friends and acquaintances (Figure S10).

**Figure 8.**
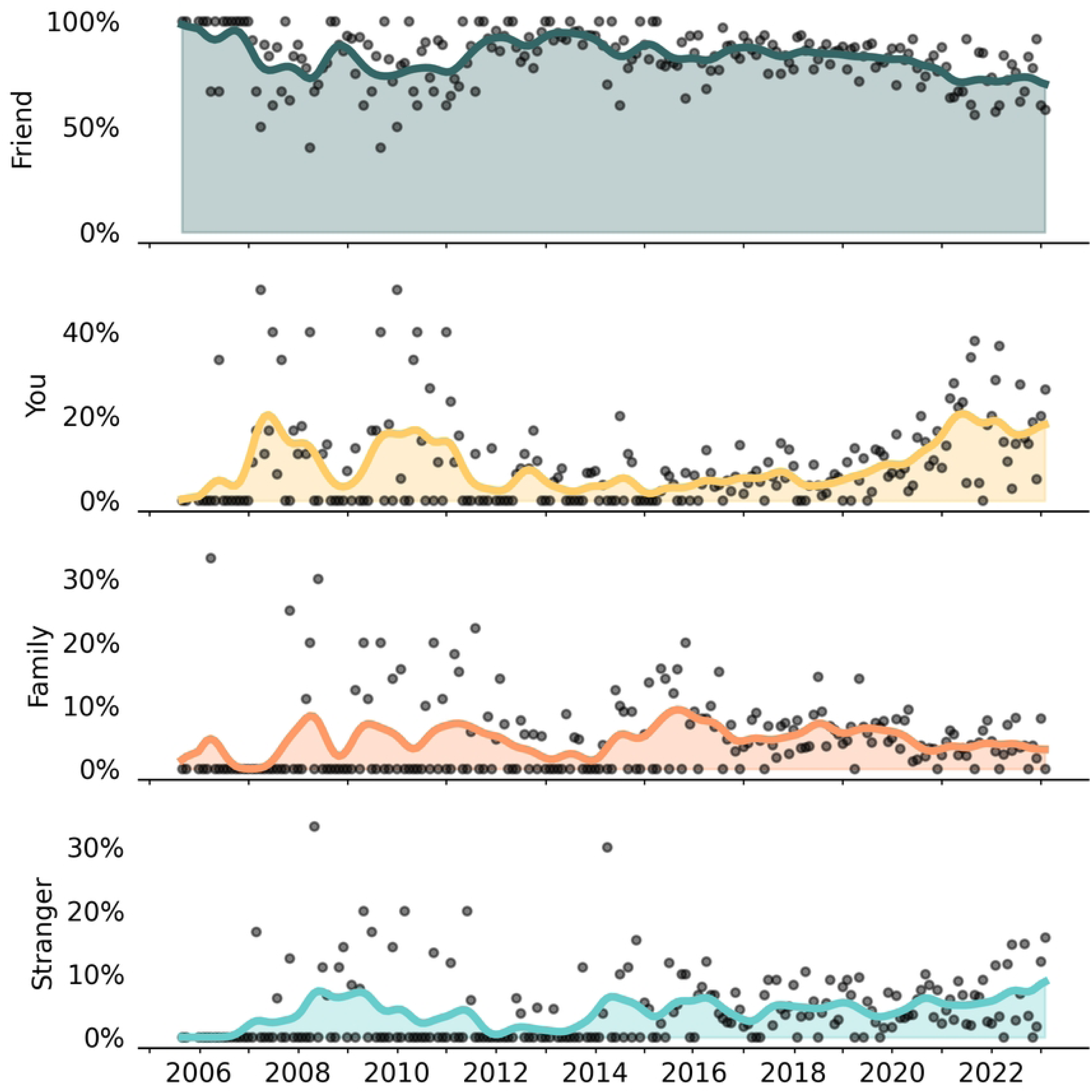
Person to whom naloxone was administered, monthly proportion of ORE reports. Dots represent monthly proportions for each exclusive category. Precents are based on total ORE reports as the denominator. The majority of naloxone administrations were performed on friends or acquaintances of the person reporting the overdose response event. “You” refers to the overdose having been experienced by the person who was reporting the overdose. Study dates: August 2005 to January 2023.

Naloxone was used on the person reporting the ORE (“you”) in only 8.9% of cases. In these cases, the person who had been prescribed naloxone was also the person who had experienced the overdose and was self-reporting the ORE. Self-reported OREs were 7.6% in the first two years of program operations and had continued to decline until May 2018 after which they approximately doubled (Table 2, Figure S9). In the last 6 months of the observation period, the person reporting the ORE was also the person who experienced the overdose in 16.3% of reports. Interestingly, the percent of OREs on the reporter themselves was twice as high for nasal sprays (16.9%) than for injectable (8.5% 10 mL vial, 5.9% 1 mL vial) formulations.

Naloxone was administered on family and strangers rarely. OREs on family members were 4.8% overall (Table 1, Figure S11). In segmented regression, family OREs climbed gradually, but July 2018 emerged as a possible change point after which OREs of family members declined. Use on family members did not differ by formulation type (range across formulations: 4.4% to 5.3%), suggesting that both intramuscular and nasal formulations were equally likely to be used on family.

During the first two years of operations, naloxone administration to strangers was less than 1% but increased to 7.9% of ORE reports in the last 6 months of observation. Cumulatively, there were 239 (4.4%) total OREs on strangers. No change point was detected in segmented regression models. The nasal spray was slightly more likely to be used on strangers (5.7%, n=78) than the 1 mL vial (4.2%, n=128), and ahead of the 10 mL vial (2.7%, n=23) distributed in early years.

#### Programmatic Context

Prevention Point Pittsburgh staff offered a plausible explanation for the increase in “you” as the person who had been revived from overdose with naloxone. In the latter two years of the observation period, a community outreach worker had been specifically engaging with “trap houses” where people congregated to use drugs [55,56]. Naloxone distribution in trap houses followed a different pattern than other types of community distribution, in what amounted to active case finding of people who had already experienced an overdose. The outreach worker would bring naloxone for distribution to the trap house, ask if anyone had recently had an overdose reversed with naloxone, and record these reversals. This practice was important for targeting naloxone distribution to a population who had already experienced an overdose and remained at elevated risk for subsequent overdoses. The corresponding data implication was that trap house delivery resulted in more “you” reports because naloxone was being provided to people *after* experiencing an overdose that had been reversed with naloxone by a peer. This stands in contrast to other community outreach settings where naloxone was provided to people *before* they had experienced an overdose.

Program staff did not have specific contextual or programmatic explanation for the increase in administration on strangers, pointing to the small numbers and cautioning against drawing conclusions. Staff emphasized that people use whichever formulation they have on hand, and that differences in rates usage by formulation are not purely a function of device usability. At Prevention Point Pittsburgh, where both intramuscular and nasal naloxone are equally available and participants are given the choice of which naloxone to take with them, many participants select intramuscular formulations if they believe *themselves* to be at risk of an overdose, and conversely, choose the nasal formulation if they expect to administer it *on acquaintances*.

Therefore, when a stranger is encountered in an unresponsive state, participants administer whichever formulation of naloxone they are carrying at the time. Staff emphasized the parity between injectable and nasal formulations, in terms of usability and preference, as reported by the participants; each has its place in community-based overdose response and there was no strong preference for the nasal spray just based on its form factor.

### Naloxone Doses Administered

#### Quantitative Results

The cumulative number of naloxone doses administered was 8,756, leading to 5,521 ORE reports, Table 3. The frequency of administered dosage forms was: 52% as 1 mL vials (n=4,551.25 doses), 25.3% as 4 mg nasal sprays (n=2,216 doses), and 18% as 10 mL vials (n=1,555.25 doses). Multiple forms of naloxone were administered in 151 ORE reports (4.7%, n=408.5 doses, Table 3).

**Table 3.** Distribution, overdose responses, and dosing by naloxone formulation.

| Naloxone Formulation | Dispensed by Prevention Point |  |
| --- | --- | --- |
|  | Units | Doses |
| 1 mL vial | 35,715 vials | 35,715 |
| Nasal spray (4 mg) | 8,032 two-packs | 16,063 |
| 10 mL vial | 1,842 vials | 18,420 |
| Autoinjector | 18 two-packs | 36 |
| Nasal adaptor |  | 0 |
| Multiple forms |  | . |
| Missing |  | 0 |
| Totals |  | 70,234 |

**OVERDOSE RESPONSE EVENTS**
|  | Reversals | % | Utilization: ORE per Unit or Dose |  |
| --- | --- | --- | --- | --- |
|  |  |  | By Units | By Dose |
| 1 mL vial | 3,082 | 56 | 8.6% | 8.63% |
| Nasal spray (4 mg) | 1,373 | 25 | 17.1% | 8.55% |
| 10 mL vial | 880 | 16 | 47.8% | 4.78% |
| Autoinjector | 14 | 0 | 77.8% | 38.9% |
| Nasal adaptor | 1 | 0 |  |  |
| Multiple forms | 151 | 3 |  |  |
| Missing | 20 | 0 |  |  |
| Totals | 5,521 | 100 |  |  |

**DOSES ADMINISTERED**
|  | Doses per ORE |  |  |  |  |  |
| --- | --- | --- | --- | --- | --- | --- |
|  | Doses | Mean | 95% CI | Relative Dose | 95% CI | Wald Test |
| 1 mL vial | 4,551.25 | 1.51 | 1.48, 1.54 | ref |  |  |
| Nasal spray (4 mg) | 2,216 | 1.63 | 1.58, 1.68 | +7.5% | 3.7%, 11.5% | $\chi^2=3.9$; $p<0.001$ |
| 10 mL vial | 1,555.25 | 1.85 | 1.75, 1.94 | +22.1% | 15.6%, 28.9% | $\chi^2=7.1$; $p<0.001$ |
| Autoinjector | 23 | 1.64 | 1.20, 2.08 | +8.6% | -16.2%, 40.7% | $\chi^2=0.6$; $p=0.53$ |
| Nasal adaptor | 2 | 2.00 | | +32.2% | 29.7%, 34.7% | $\chi^2=28.8$; $p<0.001$ |
| Multiple forms | 408.5 | 2.76 | 2.58, 2.94 | +82.4% | 70.3%, 95.4% | $\chi^2=17.1$; $p<0.001$ |
| Missing | 153 |  |  |  |  |  |
| Totals | 8,756 |  |  |  |  |  |

Poisson regression, with $\chi^2$ -scaled residuals, and robust variance estimator, using complete case data ( $n=5,375$ ). 1 mL vials used as referent group.
Model-based Wald $\chi^2$ test, df 4
\* Estimated under the assumption that doses administered were dispensed by Prevention Point Pittsburgh. As the presence of the nasal adaptor report suggests, some participants may have obtained naloxone from other sources, such as pharmacy or other harm reduction programs, and would not be accounted for in the denominator.

Cumulative utilization rates are a quantification of the number of dispensed doses used in OREs. Dose-standardized utilization rates were similar for the 1 mL vial (8.6%) and nasal spray (8.5%), and 4.8% for the 10 mL vial (Table 3).

The cumulative arithmetic average number of naloxone doses per overdose response event was 1.63 (95% CI: 1.60, 1.65), and the geometric mean was 1.44 (95% CI: 1.42, 1.46). Time trends presented in Figure 9 show two distinct patterns. The 1 mL vial and 4 mg nasal spray were used mostly as one or two doses (roughly bimodal), whereas the 10 mL vial and multiple forms represent more continuous distributions, including titrated fractional doses and administration of less than one full labeled dose to achieve reversal. When multiple forms of naloxone were administered, the number of total doses was greater (median of two versus median of one) than single formulation administrations.

**Figure 9.**
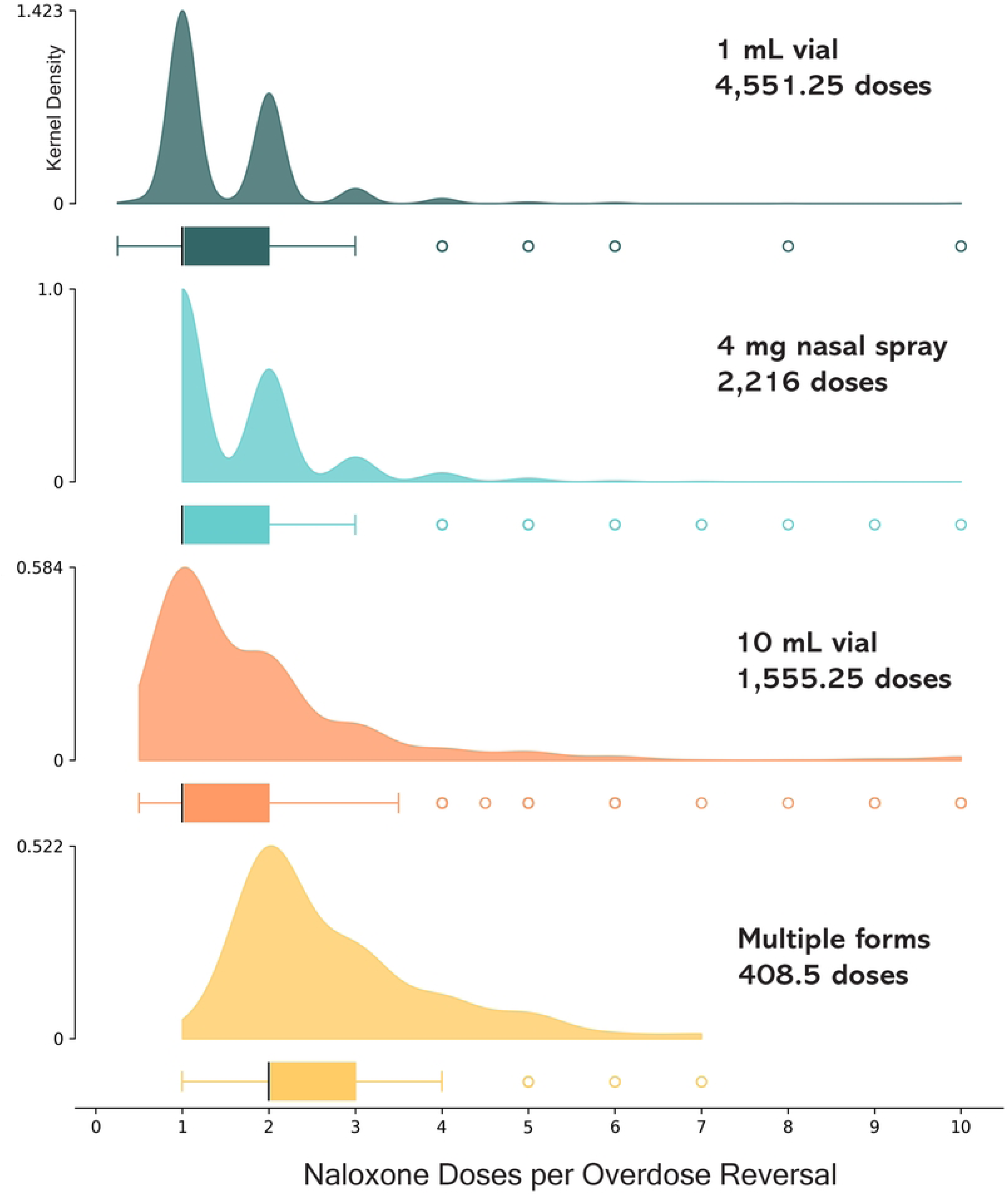
Number of naloxone doses administered per overdose response event. Average number of naloxone doses administered per overdose response event are presented with the vertical axis, smoothed using Gaussian kernel density estimators. For 1 mL and 10 mL vials, fractional dosing was observed, with less than a single full dose (1 mL of 0.4 mg/mL) being administered; this was not possible for the nasal spray, resulting in the visible left-truncation of the kernel density plot. Box plots below each graph show median and interquartile range are presented horizontally; circles represent outlier observations. Study dates: August 2005 to January 2023.

The average doses per overdose response event was lowest for 1 mL vial with 1.51 doses (95% CI: 1.48, 1.54), Table 3. Using 1 mL vials as a reference group, the average number of doses per ORE was 7.5% higher (95% CI: 3.7%, 11.5% higher, *χ*^2^=3.9, Wald p<0.001) for the nasal spray with 1.63 doses per ORE (95% CI: 1.58, 1.68). For the 10 mL vial, doses were 22.1% (95% CI: 15.6%, 28.9% higher doses, *χ* ^2^=7.1, p<0.001) higher than the 1 mL vial with 1.85 doses per ORE (95% CI: 1.75, 1.94). When multiple forms were used, the average doses were 2.76 doses per ORE (95% CI: 2.58, 2.94).

The overall rate of naloxone doses per overdose response event in Pittsburgh remained stable over a 17-year period (Figure 10). Segmented regression did not yield any statistically verifiable change points (*χ* ^2^= 0.28, 3 df, p=0.60; Table 2) during the 17-year observation period of the number of doses per overdose response event (Figure S13).

**Figure 10.**
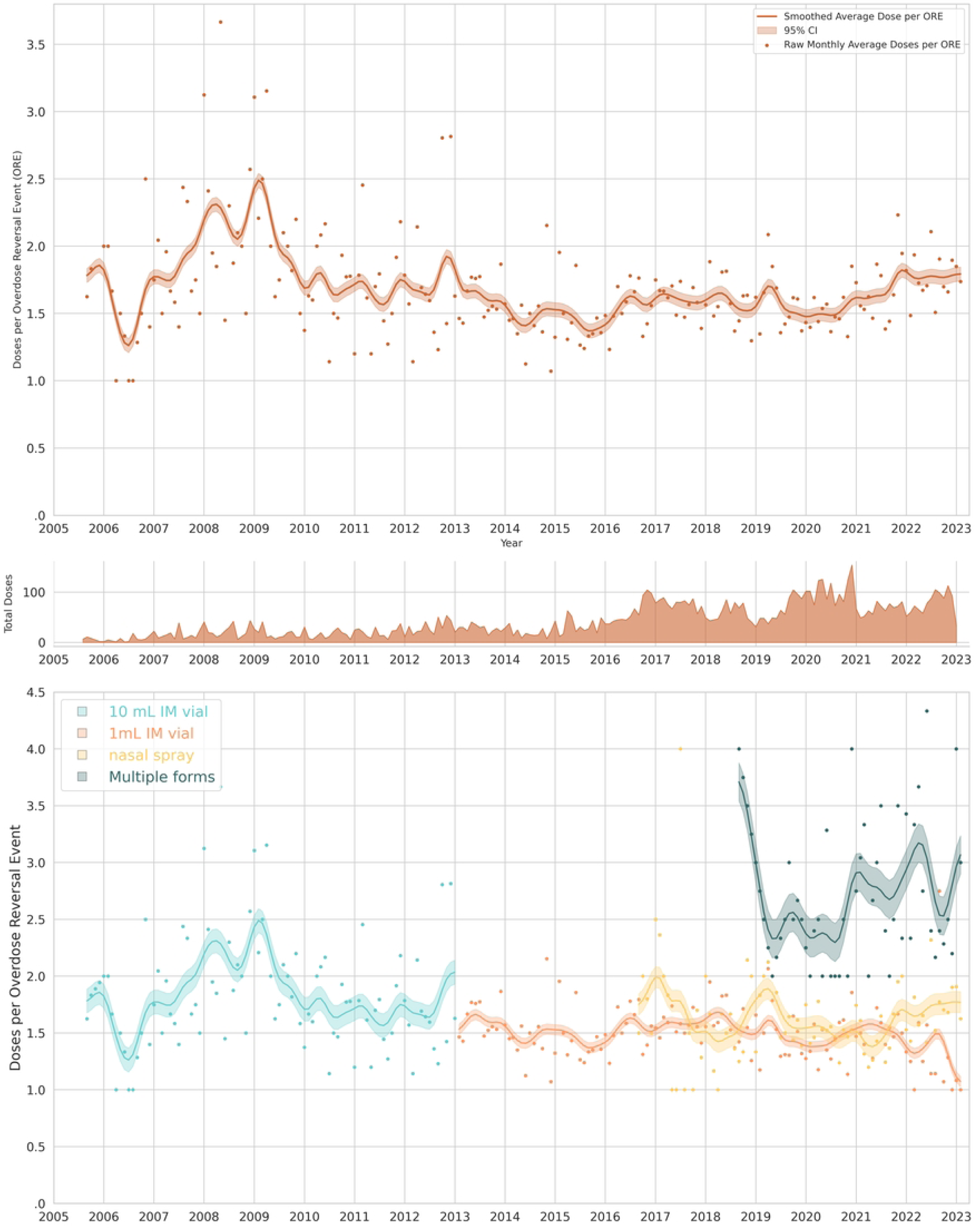
Time trends in naloxone doses per overdose response event, cumulatively and by formulation. Average number of naloxone doses administered per overdose response event for all formulations are presented in the top frame. In the bottom frame the same data are broken out by the three dominant formulations. The vertical axis corresponds to average monthly dose per overdose response event, with dots representing the raw monthly average, and the trend lines depicting the Gaussian-smoothed rolling three-month rolling average. Shaded fill areas represent the 95% confidence interval of the smoothed mean doses per overdose response event per month. Study dates: July 2005 to January 2023.

#### Programmatic Context

Program staff described that with the 10 mL vial it was easy for people to administer additional doses, and that unit dosing with the 1 mL vial and nasal spray reduced this behavior. In instances where many doses and/or multiple forms had been administered, program staff described that it was common for people to receive additional doses of naloxone after police or paramedics arrived, even if the person was already revived and was breathing. Therefore, extreme numbers of reported doses administered may reflect circumstances beyond the reporter’s control, and doses not dispensed by Prevention Point Pittsburgh. Dose titration was also identified by program staff and the rest of the study team as an important phenomenon for exploration, and one that had not been previously characterized in scientific publications.

Program staff believed that titration would be associated with fewer or less severe withdrawal-related adverse events, and this hypothesis was the basis for subsequent investigation.

### Response Behavior: Rescue Breathing

#### Quantitative Results

The percent of overdose response events in which rescue breathing was reported was 43.8% (n=2,419 out of 5,521), Table 1. Segmented regression identified January 2014 (Figure 11) as a possible changepoint after which the percent of ORE with rescue breathing declined (Figure S14). In the last 6 months of observation, 34% of overdose response events mentioned rescue breathing, down from 52% in the first 2 years of the observation period (Figure 11). The COVID-19 pandemic did not appreciably accelerate the long-term temporal trend of declining rescue breathing during overdose response events. Segmented regression with an imposed breakpoint in March 2020 did not produce a meaningful statistical association, meaning that rescue breathing continued to decline at a linear decay trajectory consistent with the immediately preceding time period (Figure S14), independent of the pandemic.

**Figure 11.**
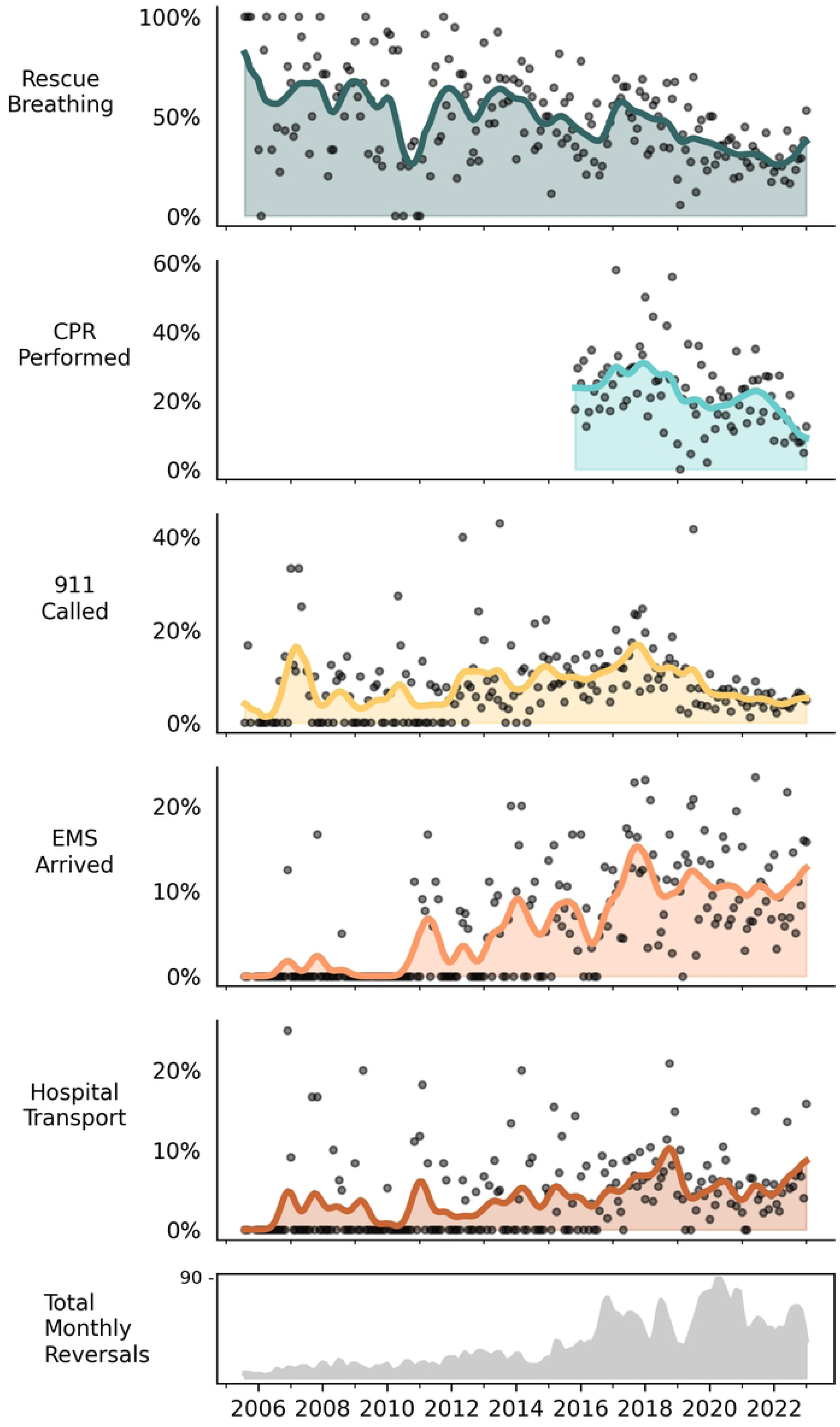
Proportion of naloxone administration reports in which response behaviors were reported. Smoothed time series depicting the percent of overdose response events in which response behaviors were reported. The bottom panel shows monthly volume of total overdose response events (ORE). Study dates: August 2005 to January 2023.

#### Programmatic Context

During the study period, Prevention Point Pittsburgh trainings recommended rescue breathing during an overdose response event. However, this recommendation was not made strongly during the height of the COVID-19 pandemic, but was re-emphasized in trainings starting in 2023, after xylazine started appearing in the local unregulated opioid supply in 2022 [57].

Program staff also noted that while rescue breathing was taught at the initial trainings, it was not reinforced during refill encounters, with one exception: If participants reported needing to use more than 2 doses of naloxone, they were counselled to use rescue breathing and count respiration in future reversals. Therefore, the slight uptick in rescue breathing in 2022 may have been influenced by this directive. In the same year, xylazine started to appear alongside fentanyl in the local unregulated drug supply, complicating reversals because the person who had overdosed may not become responsive to stimuli upon naloxone administration, even if respiration was adequately restored. It was not until March 2023, beyond the end of observation, that Prevention Point Pittsburgh formalized the recommendation to reemphasize counting breaths after naloxone administration (and provide rescue breathing if needed) to participants via handouts and flyers in the context of xylazine; the intent was to dissuade unnecessarily high doses of naloxone being administered if the participant was breathing adequately but did not become immediately reanimated. This was also contemporaneous with the start of disposable xylazine test strip distribution. While this programmatic evolution was not formalized until after the end of the observation period, Prevention Point Pittsburgh noted that the same advice had been delivered to participants in less systematic ways in the months prior. Staff also pointed out that the current over-the-counter label [43] for naloxone nasal sprays does not include instructions for rescue breathing but that they provide the instructions themselves.

### Response Behavior: Chest Compressions

#### Quantitative Results

Data on chest compressions (commonly called “CPR” for cardio-pulmonary resuscitation) were recorded from 2016 to 2023. Chest compressions were reported to have been conducted in only 23.7% of OREs (n=711 out of 3,004 reports). Segmented regression identified February 2017 as a changepoint after which chest compressions declined (Table 2, Figure 11)

#### Programmatic Context

Prevention Point Pittsburgh staff pointed out that in 2016, the New York State Health Department convened a working group of physicians and scientists to establish the necessity of recommending CPR for opioid overdose reversal. At the time, CPR was recommended widely in Canada but not by the World Health Organization, and within the US instructions were inconsistent. The onset of data collection at Prevention Point was intended to inform this debate. The medical consensus of the New York State working group was that chest compressions were not required [58]. As a result, Prevention Point Pittsburgh modified recommendations to participants de-emphasizing chest compressions, which had been part of the original training. They attribute the decline in CPR in 2017 to be related to this programmatic shift.

### Response Behavior: Calling 911

#### Quantitative Results

Overall, 911 was called in 16.4% (n=903 out of 5,521) of ORE reports (Table 1, Figure 11). When 911 was called, EMS arrived half the time, or 49.2% of reports (n=444 out of 903, with 0.3% missing). Segmented regression identified December 2017 as a changepoint after which calls to 911 decreased considerably; during the last 6 months of the observation period, 911 was called in only 5.4% of OREs.

#### Programmatic Context

Calling emergency medical services (EMS, i.e., 911) was universally recommended in trainings by Prevention Point Pittsburgh. However, fear of criminal prosecution is expected to have dampened the likelihood of this behavior. The Pennsylvania “Good Samaritan” Act 139 was enacted in December 2014, offering limited criminal immunity for minor drug possession charges when calling EMS during an overdose [59]. In spline regression, comparing before and after the law change (Figure S17), there appears to have been a transitory increase in calls to 911, which was not sustained over time. Program staff also stated that people who do not use drugs are more likely to call 911, and increases in dispensing naloxone outside of networks of people who use drugs in the years after state legislation enactment could have contributed to the transitory increase in 911 calls.

Program staff offered explanations to account for the surprisingly low rate of EMS arrival, based on conversations during the ORE report intake that were not quantified on forms. The common theme was wanting to avoid encountering first responders (which could include police) unless absolutely necessary, especially in the context of drug-induced homicide laws as documented elsewhere [60]. Successful reversals may have resulted in a follow-up call to 911 stating that EMS were no longer required. The reporter may have called EMS, administered naloxone, and then left the scene, and therefore may not know for certain if EMS arrived. For these reasons, program staff cautioned in the interpretation of the seemingly low rate of EMS arrival in these data.

### Response Behavior: Hospital Transport

#### Quantitative Results

Among the 5,521 ORE reports, hospital transport was reported in 5.0% (n=274) of OREs. No temporal changepoint was detected in segmented regression, Table 2. Visual inspection of the time trend revealed heavy concentration of zeros (e.g., no hospital transport) in the first 11 years of the observation period (Figure 11). The last six months of the observation period suggested an uptick, with 7.7% (95% CI: 3.3%, 12.0%) of OREs having had hospital transport.

#### Programmatic Context

Empirical data show that hospital transport had a sudden peak in 2017 [61]; program staff pointed out that this was the year with peak overdose deaths in Allegheny County, as well being contemporaneous with the brief emergence of carfentanil in the unregulated drug supply.

Additionally, based on interviews during ORE reports, program staff suggested that emergence of xylazine in 2022 may have led to more hospital transport due to lack of reanimation.

### Adverse Events

Granular information on specific adverse events (AEs) was recorded systematically starting in August 2016 and are thus available for 4,606 ORE reports, which constitutes the denominator for rate calculations. Any adverse event was noted in 31.4% (n=1,446/4,606) of OREs. Overall, emesis (vomiting), anger, and “feeling sick” were the most commonly reported adverse events. Annualized time trends in AE counts and rates per OREs are displayed in Figure 12 for the 6 calendar years (2017-22) with complete reporting. The two most common AEs (Table 4) described in free text notes were confusion (n=11) and diarrhea (n=4), which only were reported with the 1 mL vial. There was no indication in free text fields for wooden chest syndrome, muscle rigidity, laryngospasm, or pulmonary edema, which are other adverse events of concern with illicitly manufactured synthetic opioids.

**Figure 12.**
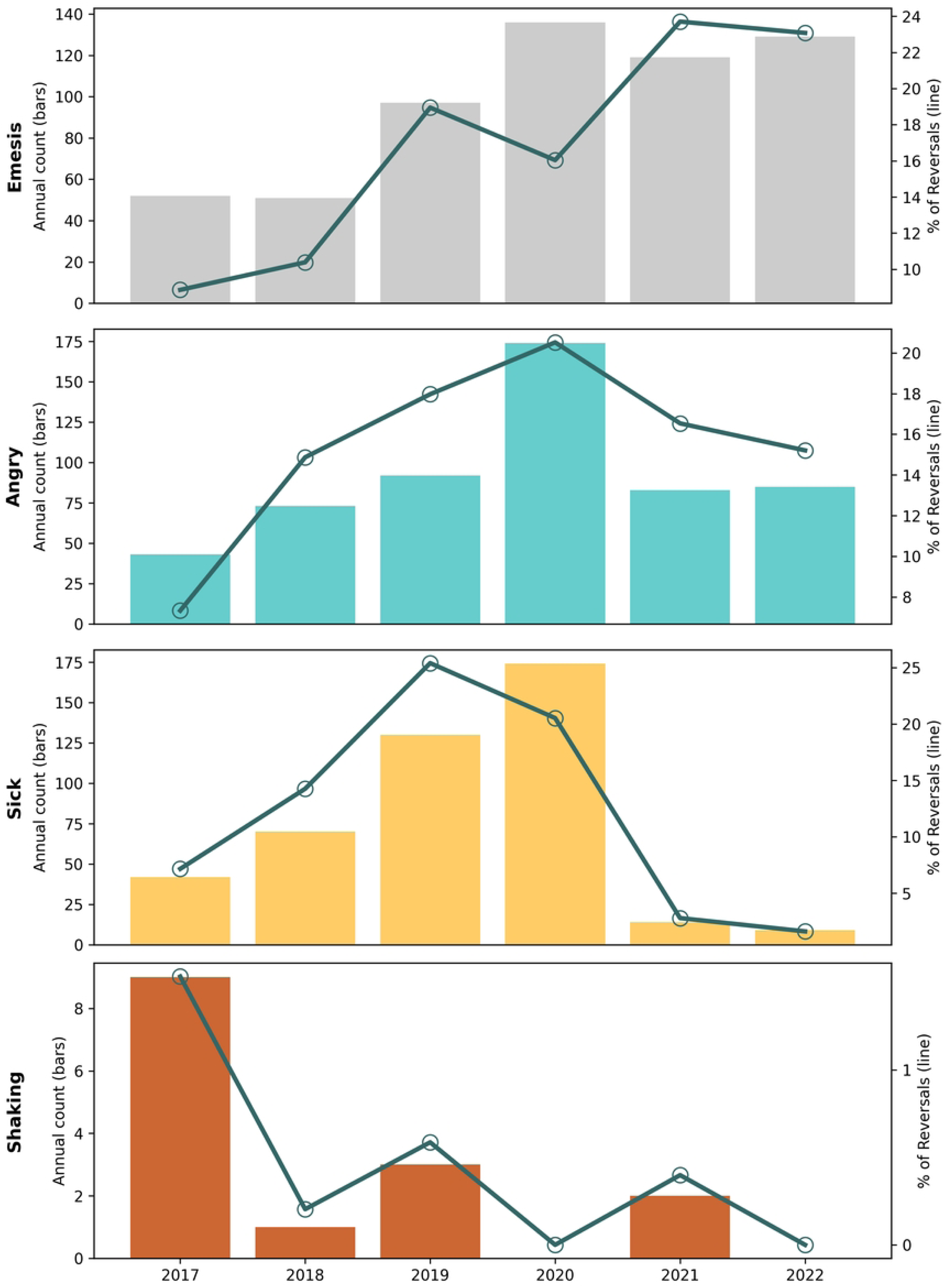
Annual counts and percents of adverse events following naloxone administration. Vertical bars are counts of adverse event reports by year. Green lines are percents of overdose response events annually (N=4,606 total), enumerated on the right vertical axis.

**Table 4.** Adverse events reported after naloxone administration, by formulation and dose, August 2016 to January 2023.

|  |  | Naloxone Formulation |  |  |  | Doses Administered |  |  |  |
| --- | --- | --- | --- | --- | --- | --- | --- | --- | --- |
|  |  | Total | 1 mL vial | Nasal 4 mg | Multiple Forms | Wald Test* | Average Doses<br>(95% CI) | Cases<br>>1<br>Dose | Percent<br>of Cases<br>> 1 Dose |
|  | All Reversals | 4,606 | 3,082 | 1,373 | 151 |  | 1.59 (1.56, 1.61) | 1,972 | 43.6% |
|  | Reversals with any AE recorded below | 1,446 | 820 | 557 | 69 |  | 1.66 (1.62, 1.71) | 685 | 47.8% |
| Emesis | Cases | 607 | 296 | 284 | 31 | x2 108,<br>p<0.001 | 1.73 (1.65, 1.81) | 308 | 50.7% |
|  | (Percent) or Rate per 1,000 Reversals | (13.2%) | 96.2 | 207.6 | 205.3 |  |  |  |  |
|  | Rate Difference per 1,000 reversals<br>relative to 1 mL vial, adjusted for dose |  | ref | +106 | +90.3 |  |  |  |  |
|  | Rate difference 95% CI |  |  | +82.0, +130 | +22.6, +158 |  |  |  |  |
| Angry | Cases | 546 | 314 | 206 | 31 | x2 31,<br>p<0.001 | 1.61 (1.53, 1.68) | 237 | 43.4% |
|  | (Percent) or Rate per 1,000 Reversals | (11.9%) | 102.1 | 150.6 | 205.3 |  |  |  |  |
|  | Rate Difference per 1,000 reversals<br>relative to 1 mL vial, adjusted for dose |  | ref | +48.5 | +103 |  |  |  |  |
|  | Rate difference 95% CI |  |  | +47.8, +49.2 | +101, +105 |  |  |  |  |
| Felt Sick | Cases | 440 | 291 | 136 | 18 | x2 1.1,<br>p=0.56 | 1.58 (1.50, 1.67) | 186 | 42.3% |
|  | (Percent) or Rate per 1,000 Reversals | (9.6%) | 94.6 | 99.4 | 119.2 |  |  |  |  |
|  | Rate Difference per 1,000 reversals<br>relative to 1 mL vial, adjusted for dose |  | ref | +4.8 | +24.6 |  |  |  |  |
|  | Rate difference 95% CI |  |  | +4.2, +5.4 | +22.8, +26.4 |  |  |  |  |
| **Death** | Cases | 47 | 24 | 20 | 3 | x2 5.6, exact<br>p=0.043 | 1.94 (1.63, 2.24) | 29 | 61.7% |
|  | (Percent) or Rate per 1,000 Reversals | (1.02%) | 7.8 | 14.6 | 2.0 |  |  |  |  |
|  | Rate Difference per 1,000 reversals<br>relative to 1 mL vial, adjusted for dose |  | ref | +4.5 | +8.4 |  |  |  |  |
|  | Rate difference 95% CI |  |  | -2.6, +11.6 | -14.9, +31.6 |  |  |  |  |
| **Shaking** | Cases | 33 | 27 | 5 | 1 | x2 10.5,<br>exact<br>p=0.005 | 1.97 (1.50, 2.43) | 21 | 63.4% |
|  | (Percent) or Rate per 1,000 Reversals | (0.7%) | 8.8 | 3.7 | 6.6 |  |  |  |  |
|  | Rate Difference per 1,000 reversals<br>relative to 1 mL vial |  | ref | -5.1 | -2.1 |  |  |  |  |
|  | Rate difference 95% CI |  |  | -5.0, -5.3 | -1.7, -2.6 |  |  |  |  |
| **Confusion\*\*** | Cases | 11 | 11 | 0 | 0 |  | 2.36 (1.40, 3.32) | 8 | 72.7% |
|  | Rate per 1,000 Reversals |  | 3.6 |  |  |  |  |  |  |
| **Diarrhea\*\*** | Cases | 4 | 4 | 0 | 0 |  | 1.25 (0.45, 2.0) | 1 | 25.0% |
|  | Rate per 1,000 Reversals |  | 1.3 |  |  |  |  |  |  |

\* Model-based Wald chi-square test with 2 df. Fisher's Exact test was used if any cell count was less than 10.
\*\* Interpret with caution. Derived from free text notes, not recorded on structured form.

OREs in which adverse events were reported had slightly higher average naloxone doses 1.66 (95% CI: 1.62, 1.71) compared with all OREs 1.59 (95% CI: 1.56, 1.61). More than one dose of naloxone was associated with higher incidence of any reported adverse event, 47.3% of OREs (n=685/1,446), compared to 40.7% (n=1,287/3,160) of OREs when one or fewer doses were administered (Wald *χ* ^2^=17.8, 1 df, p<0.001), a rate difference of 6.6 per 100 OREs (95% CI: 3.5, 9.7).

### Adverse Events: Emesis

#### Quantitative Results

Emesis (vomiting or “puking”) was reported in 13.2% (n=607) of OREs, but differed (Wald x^2^ 108, 2 df, *p*<0.001) considerably by formulation. OREs using the 4 mg nasal spray were twice as likely to result in emesis compared to the 1 mL vial (20.8% versus 9.6%), Table 4. After adjusting for doses administered, there were 106 (95% CI: 82, 130) more emesis events per 1,000 OREs with the 4 mg nasal spray than the 1 mL vial at 0.4 mg/mL.

In both absolute and relative (to OREs) rates, Figure 12, emesis increased as a reported adverse event from 2017 to 2022. In the era when only 1 mL vials were distributed, about 16% of ORE involved emesis, going up to about 23% when naloxone distribution included vials and 4 mg nasal spray both, contemporaneous also with the advent of illicitly manufactured fentanyl.

#### Programmatic Context

Prevention Point Pittsburgh staff noted that emesis was the most objectively observable adverse event, and that it would be more likely to be reported than other more subjective adverse events, impacting interpretation of relative prevalence between AEs. But they did not identify a reason why reporting of emesis would be different between formulations, providing credence to inter-formulation comparisons.

### Adverse Events: Anger

#### Quantitative Results

Anger was reported in 11.9% of OREs (n=546/4606) and differed by formulation (Wald *χ* ^2^ 108, 2 df, *p*<0.001). The 1 mL vial was associated with fewer angry AE reports, at 10.2%, compared to 15.1% of OREs with the 4 mg nasal spray, Table 4. After adjusting for doses administered, per 1000 OREs there were 48.5 (95% CI: 47.8, 49.2) additional cases of anger after administration of the nasal spray.

In both absolute and relative measures, anger after naloxone administration was most reported in 2020, during the phases of COVID-19 pandemic isolation. However, even though 4 mg nasal spray supplanted 1 mL vials during a sudden injectable naloxone supply shortfall in 2021-22 (Figure 5), reports of anger after naloxone administration returned to pre-pandemic levels despite the formulation change. This shortfall was the result of manufacturing difficulties at the single manufacturer that supplied the Naloxone Buyers Club, resulting in nationwide lack of availability for harm reduction programs [62].

#### Programmatic Context

Prevention Point Pittsburgh staff cautioned that if someone was vomiting, they may not be able to simultaneously express anger, and therefore the reported numbers are likely an undercount of experience. Perceptions of what constitutes anger could also be subjective, and in very rare cases extreme, with confrontational action against the person reversing the overdose. But they did not identify reasons why reporting would be different by formulation.

### Adverse Events: “Felt Sick”

#### Quantitative Results

“Felt sick” was understood by participants to mean “dopesick” from precipitated opioid withdrawal. Cumulatively, 440 cases of “feeling sick” were reported, in 9.6% of OREs (Table 4). The incidence rate difference between formulations was negligible.

Adverse events where the recipient “felt sick” were highest in 2019, in nearly 25% of OREs, but highest in terms of absolute number (about 175 per year) in 2020, Figure 12. There was a drop in reports of “feeling sick” in 2021-2.

#### Programmatic Context

Prevention Point Pittsburgh staff did not have a specific attribution to explain temporal variation in feeling sick. However, they pointed out that feeling dopesick was a subjective experience that may not be entirely or objectively observable by the person who administered naloxone and was providing the ORE report.

### Adverse Events: Shaking

#### Quantitative Results

Shaking was systematically collected, and was reported in n=33 OREs, and differentially by formulation: n=27 cases using the 1 mL vial, n=5 with nasal alone, and n=1 with multiple forms (Fisher’s Exact 10.8, 2 df, *p*<0.005). The incidence rate difference of −5.1 cases (95% CI: −5.0, - 5.3) per 1,000 OREs slightly favored the nasal spray over the 1 mL vial. Shaking was mostly reported in 2017, Figure 12, with 9 cases, and three or fewer in subsequent years.

#### Programmatic Context

Shaking was interpreted by Prevention Point Staff to be a sign of opioid withdrawal, as opposed to seizures. No plausible explanation for this phenomenon was offered by Prevention Point Pittsburgh staff.

### Adverse Events: “Was Okay” versus Death

#### Quantitative Results

Participants presenting for refills had been asked if the person on whom naloxone was administered “was okay” after the ORE, to the best of their knowledge, since the start of study observation. This category is conceptually understood to encompass survivorship, even if hospital transport or adverse events occurred, and serves as a contextual adjunct to deaths. Cumulatively, in 5,449 out of 5,521 reports (98.7%) from 2005 to 2023, the participant felt confident enough to respond. The person on whom naloxone had been used was judged to be “okay” 98.0% of the time (n=5,340/5,449 among the three dominant naloxone formulations).

Differences by formulation were observed (Pearson *χ* ^2^ 32.7, 3 df, p<0.001), but were small: 98.5% okay with 1 mL vial, 99.2% with 10 mL vial, 96.4% with 4 mg nasal spray, and 95.4% with multiple forms.

Death as an adverse event was collected from 2016 onwards. There were 47 out of 4,606 reports (1.0%) of death following naloxone administration.

#### Programmatic Context

Program staff had strong cautions about interpretation of deaths following naloxone administration. The death case review provides additional context below. In addition, program staff felt that as the volume of naloxone distributed has increased within networks of people using drugs together, there should be increased likelihood of someone at the scene carrying naloxone. Therefore, they interpreted deaths following naloxone administration as an indirect indicator of people who had been using drugs alone. Forced isolation during the first year of the COVID pandemic provided an opportunity to test this hypothesis, as described below.

### Death Case Review

#### Quantitative Results

From 2020 through 2022, encounter notes with contextual information were available for 22 out of 23 deaths. In 18 cases the person was found “too late” after death for naloxone to be effective, for example the morning after an overdose that had likely occurred the previous evening. In three cases, paramedics told the reporter that there was an alternative cause of death other than overdose. In one instance, a reporter said that police had not allowed her to administer the naloxone that she was carrying, and the person died.

Quantitative AE data allowed empiric corroboration that deaths increased during the height of pandemic isolation. Most years, 5 or fewer deaths were reported, Figure 13. However, in 2020, the annual reports tripled in both absolute and relative terms. This single-year increase paralleled the time-trend with AEs for anger, but with much lower sample size.

**Figure 13.**
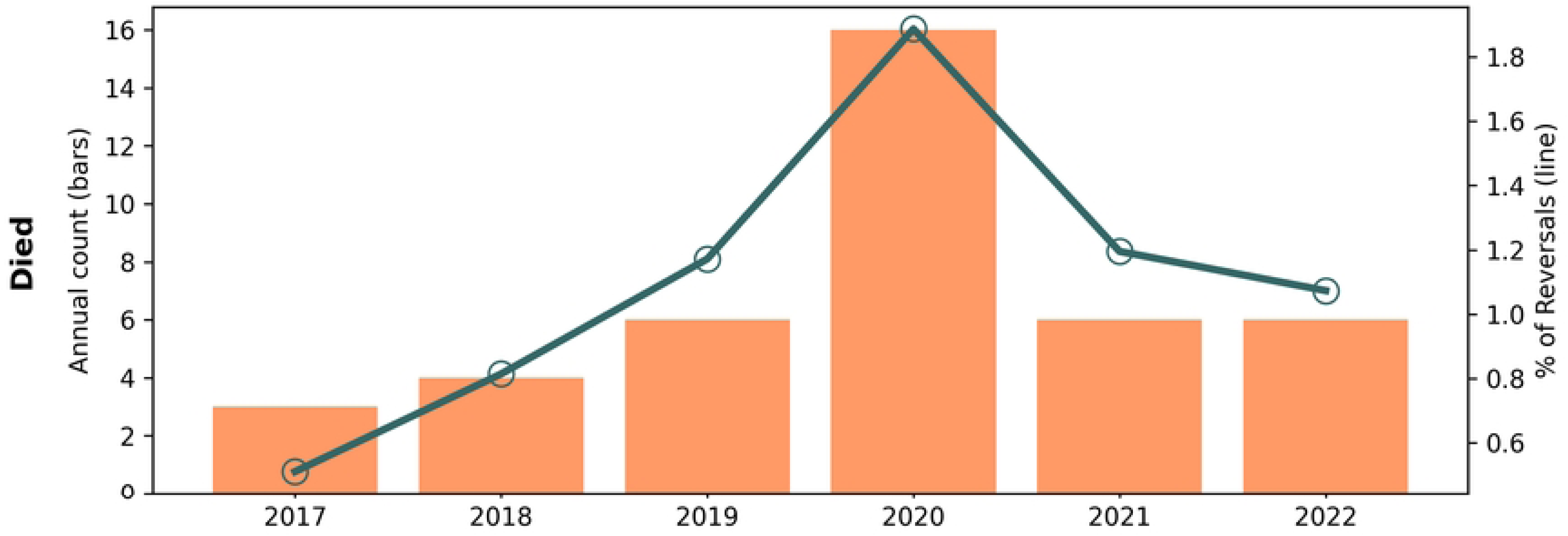
Annual counts and percents of deaths reported. Vertical bars are counts of adverse event reports by year. Green lines are percents of overdose response events annually (N=4,606 total), enumerated on the right vertical axis.

#### Programmatic Context

Program staff posited that, under the assumption that using drugs alone was more common during forced social isolation and quarantine periods of the first year of the COVID-19 pandemic, people who had overdosed may have been less likely to have been found for timely action. Alternatively, additional respiratory compromise stemming from viral infection could be a plausible contributory cause. Overwhelmed EMS during the early pandemic could also in part explain the single year increase in reported deaths. Program staff emphasized that they did not experience a drop in service provision during COVID and increased how much supplies were distributed per visit to reduce the need for return visits, as well as providing additional services (food, clothing, etc.) to meet community needs.

### Adverse Events and Dose Titration

During the period where AE information was being systematically recorded (2016 onwards), the 1 mL vial offers a unique opportunity to conceptually explore dose-response between naloxone dose and adverse events. The expectation was that naloxone dose titration would result in fewer adverse events. Titration was defined as reporting fractional dosing (n=117 OREs), and non-titration was defined as integer doses of naloxone administered (n=2,953 OREs). This imperfect exploratory measure deserves reiteration of the caveat for misclassification: Two half doses administered separately would be reported as 1.0 dose, but in this simple metric would be misclassified integer dosing. This is expected to bias towards the null.

Titration of naloxone was recorded in only 4.0% of OREs, but titration of naloxone showed favorable incidence rate ratios (IRR) for the three most common adverse events (Table S2). Titration was associated with less emesis IRR=0.26 (95% CI: 0.084, 0.80; Wald *χ* ^2^ 2.36, *p*=0.018), less anger IRR=0.081 (95% CI: 0.011, 0.57; Wald *χ* ^2^ 2.5, p=0.012), and less “feeling sick” IRR=0.44 (95% CI: 0.186, 1.051; Wald X^2^ 3.8, *p*=0.051). Death and shaking also showed favorable tabular distributions for titration, but model-based IRRs could not be computed due to zero cell counts.

## DISCUSSION

Community naloxone distribution at Prevention Point Pittsburgh resulted in more than five thousand reported overdose response events, over 17 years. However, previous studies of community naloxone distribution are of short duration, providing limited insight into program implementation and evolution. In addition, the preponderance of real-world data on overdose reversal with naloxone has originated from emergency medical settings and hospitals, leaving a gap in the increasingly common practice of community overdose response. Therefore, this comprehensive report includes key findings from both implementation and biomedical perspectives.

In this sample of individuals returning to a harm reduction program for naloxone refills, the majority of reported OREs indicated success (i.e., the person survived), even in recent years with fentanyl, xylazine, and methamphetamine prevalent in the local drug supply. A minority, but not a trivial proportion, had adverse events suggesting precipitated withdrawal.

### Research Questions 1 & 2: Naloxone Utilization after Law Change

State law changes at the end of 2014 were intended to remove barriers to naloxone distribution for harm reduction programs, thereby enabling the intervention to reach broader populations.

The key programmatic finding was that enabling state legislation alone was insufficient to expand services to populations at greatest risk for overdose [45]. Before the law change, which expanded legal protections and relaxed prescribing policies, Prevention Point Pittsburgh served a predominantly White network of people who used heroin, a cohort that remained steady and aged over time. While enabling legislation in 2015 led to greater naloxone dispensing volume (10.4 doses per month to 65.9 per month in the year that followed), ORE report volume did not keep apace: ORE per new participant fell from 1.46 to 0.47 in the year before versus after the law change. Demographic characteristics offer explanations. In the year after the law was enacted, new participants were more likely to be older (average age increasing from 37 to 46 years-old), more female (increasing from 35% to 58%), and family or friends of people who use drugs, a marked shift. Compared to a previous evaluation of Prevention Point data, our observations are consistent: After enabling legislation, the earlier analysis found that these demographically different new participants were only 0.04 times as likely to reverse an overdose compared to people who use drugs [16]. Instead, refocusing distribution directly to networks of people who use drugs required programmatic effort and intentionality including hiring people from target communities, providing safer smoking supplies, and starting mobile buprenorphine induction services.

During the first decade of naloxone distribution and before the 2015 enabling legislation, the most common response to whom naloxone was administered was “friend or acquaintance.” This could reflect social desirability bias due to the stigmatized nature of the intervention at the time, and fear of revealing that the participant had transferred the naloxone to someone to whom it had not been prescribed. Naloxone reversals were concentrated within social networks: 81% of OREs were performed on friends and acquaintances, not the person to whom it had been prescribed. Less than 5% was used on family members or the individual who reported the ORE. Interestingly, the percent of reversals on the reporter themselves (i.e., the person prescribed the naloxone) was twice as high for nasal sprays (16.9%) than for injectable (8.5% 10mL vial, 5.9% 1mL vial) formulations.

Monthly ORE rates (per 100 doses dispensed, or number of new participants, Figure 3) offer new possibilities for tracking the impact of policy or programmatic changes in naloxone distribution. ORE per new participants closely reflects the observations of program staff during corresponding time periods; although the metric is useful for retrospective analysis, it confirms what the program already knew from their direct care provision experience. In addition, time series of OREs per 100 doses dispensed also showed obvious peaks when there were fluctuations in the drug supply, namely the emergence of illicitly manufactured fentanyl and carfentanil. We suggest that these metrics may be useful in future epidemiologic studies and serve as a useful tool for programs to monitor. However, drawing from the research team’s national experience providing technical assistance, we also acknowledge that reversal record keeping requirements from funders can be a considerable impediment to actual service delivery. Therefore, we feel that data utility should be secondary to naloxone distribution, and the former should not impede the latter.

### Research Question 3: Racial Identity of New Participants

Addressing our third research question, as overdose death rates in underserved racialized minority communities began to increase, the program intentionally fostered new mobile outreach sites in January 2016, and started distributing safer smoking supplies that met the needs of that community. Black racial identity of new participants increased from 27% to 39%. These innovations led to significantly more naloxone distribution *and* ORE reports. However, time-series evaluation identified an inflection point in December 2020 after which the share of White participants increased. Programmatic context revealed that this was due to expansion of mobile services to additional neighborhoods to provide on-demand buprenorphine treatment for opioid use disorders, where take-home naloxone was also provided. Racial disparities in the uptake of buprenorphine services have been well-documented [63,64], and are known to have been exacerbated during the COVID pandemic [65]. These national trends and the experience at Prevention Point Pittsburgh are consistent. The findings suggest that new targeted programmatic strategies will be needed to expand medication assisted treatment to communities of color.

### Research Questions 4 & 5: Doses per Overdose Response Event

Per our fourth and fifth research questions, we found that the average doses of naloxone needed to reverse an overdose has not changed over time (segmented regression change point *p*=0.60); however, formulation effects were also observed. In the first decade of operation when overdoses were predominantly due to heroin, distribution of 10 mL naloxone vials resulted in an average of 1.85 doses per ORE. When naloxone distribution shifted to single-unit packaging, coincident with the appearance of illicitly manufactured fentanyl in the local drug supply [46], average doses were lower: 1.51 doses per ORE with the 1 mL vials, and 1.63 doses with the 4 mg nasal spray. Utilization implications for single-versus multi-dose packaging for liquid pharmaceuticals has been explored in the context of image contrast media [66] and vaccines [67], but has not been previously described for naloxone. However, studies on formulation preference between injectable and nasal forms of take-home naloxone have revealed mixed desires among the target population [68], and the current programmatic recommendation is to offer both. Programmatic context in our study revealed that preferences are not static within an individual and are related to how might be used: injectable for home and to be used on themselves, versus nasal spray for carrying in a purse to be used on others. Higher dose naloxone products were not evaluated in this study, but have been described elsewhere [69,70]. It is worth contextualizing dosing trends as the overdose epidemic evolved from one of prescription opioids to heroin, and from heroin to fentanyl – three periods defined as a “triple wave” phenomenon [1]. Using state Allegheny County inflections in fatal and non-fatal overdose to define these waves [71,72], average naloxone per ORE decreased from 1.92 (95%CI: 1.79, 2.05) in the first wave (2008-2011) to 1.57 (95% CI: 1.50, 1.64) in the second wave (2012-2015), then remained stable between the second and third (2016-2023) waves (1.60 doses, 95% CI: 1.57, 1.63). These data may be partly confounded by formulation trends discussed above; further investigation into the relationship between the drug supply and ORE behaviors is warranted.

There was also an uptick in doses per ORE administered in 2022, as xylazine entered the local drug supply [73]. Average dose of naloxone in 2021 was 1.67 (95% CI: 1.58, 1.75) increasing to 1.75 (95% CI: 1.67, 1.84) in 2022. Additional doses may have been administered because lack of reanimation even if respiration was restored. Program staff adapted to this circumstance by emphasizing counting breaths before administering more naloxone, and specifically counselling respondents who reported using more than two doses. The influence of xylazine on community-based naloxone administration practice needs further research.

### Research Question 6: Calling 911

We found that 911 was only called in 16% of ORE reports across the 17.6 year observation period. There was a transient increase in calls to 911 after enactment of the Good Samaritan law in January 2015, but by December 2017 the proportion dropped considerably, so much so that in the last 6 months of observation, less than 6% of OREs involved calling 911. There is ample evidence that people who use drugs remain fearful of arrests [74–77] when calling 911, despite the Good Samaritan law. Other possible factors that could be considered include the perception that there is no need for further treatment, fear of getting additional naloxone from uniformed first responders, stigma, cost, and wanting to use again because of withdrawal symptoms. The observed reluctance to call 911 also has implications when considering data derived from EMS and other medical encounters, given that these represent an unknown fraction of actual overdose reversals and are therefore a selected sample that may not be representative of all reversal events, in terms of formulations, doses, AEs, and outcomes.

### Research Question 7: Deaths

Research question 7, which addressed circumstances surrounding deaths, was examined using empirical findings and narrative review of reports of deaths. Most deaths (18 out of 23) occurred when the person was found “too late” to intervene. We were able to observe that deaths peaked in 2020 during the isolation phase of the COVID pandemic. Program staff emphasized that in all their years working in this program, they are not aware of any participants stating that they had administered naloxone but the person still died because they did not have enough naloxone.

There were 4.5 per 1,000 OREs more deaths reported with the nasal spray than 1 mL vial; however, strong cautions are warranted about drawing conclusions because using alone and being discovered “too late” could confound the empirical observation.

### Research Question 8: Adverse Events and Titration

Per research question 8, concerning the relationship between formulation or dose titration and adverse events, we found significant differences by formulation for the most common AEs, emesis and anger. Per 1,000 OREs, there were 106 additional reports of emesis with the 4 mg nasal spray compared to 1 mL vials; for anger, there were 48 additional reports per 1,000 OREs. Events of wooden chest syndrome and stiffness, related to synthetic opioid exposure, were not reported in this study.

### Limitations

This study has several limitations that should be considered. It represents the experience of a single harm reduction program and, per the EMI framework, study results are not generalizable to other community settings or cities. Self-reported interview data are subject to recall bias, with respondents potentially more likely to remember extreme or negative events. Participant reports were from laypersons without medical training, and reporting of adverse events may have varying accuracy. Naloxone obtained from other sources in Pittsburgh are not captured in dispensing data. While it is possible that the same overdose event may be reported by the person who administered naloxone as well as the person to whom it was administered, this is unlikely because *refills* were recorded. It was not possible to link to hospital or vital statistics in this anonymized dataset. There was no way to observe the counterfactual, namely what would have happened if the antidote had not been administered; some overdoses may have been self-resolving without naloxone administration. Despite these limitations, the detailed quantitative and programmatic context documented provide a broad historical perspective on naloxone distribution and use.

Finally, we acknowledge that record keeping of OREs has been contentious among harm reduction programs because it can place substantial administrative burden on staff that detracts from their ability to distribute naloxone and provide other direct services. By adopting the collaborative EMI process, we present an alternative model whereby important programmatic considerations and quantitative insights are given equal credence, and where the research questions are mutually agreed upon.

### Policy Implications

Our findings suggest five policy implications. First, enabling legislation to expand naloxone access is necessary but insufficient alone to reach those at highest risk for overdose. Laws and policies intended to expand community-based naloxone distribution should consider what additional practical support is required to reach the underserved. In this example from Prevention Point Pittsburgh, outreach to underserved communities and high-risk populations represent deliberate strategies that were enabled by the legislation. Diversification of harm reduction services beyond naloxone and provision of sterile injecting equipment can also increase naloxone dispensing, such as occurred in this setting with the inclusion of safer smoking supplies and mobile buprenorphine services. These program adaptations are consistent with national trends; the Centers for Disease Control & Prevention recently reported that smoking has supplanted injection as the route of administration most often implicated in overdose death [78].

Second, enabling legislation in Pennsylvania led to a decrease in naloxone being confiscated by law enforcement, but one death was reported following an incident in which someone carrying naloxone was prohibited by police from administering it. In addition, reports of excessive additional doses administered by EMS and police after successful revival by peers should be investigated. EMS policy and protocols should be re-evaluated in the context of bystander administration to ensure naloxone dosing conforms to evolving medical best practice.

Third, the Good Samaritan law appeared to have an observable but time-limited effect on increasing calls to 911. These state laws may need to be revised if they are to be more effective. The very low rate of calling 911 in recent years bears further investigation [79], such as to clarify when support services are most medically necessary, and identify social and legal barriers including the impact of drug-induced homicide laws [80]. These investigations will be crucial in the context of over-the-counter naloxone, which has explicit instructions to call 911.

Fourth, consistent with previous analyses [16,19,81], we found that 1 mL vials injected intramuscularly were effectively used in thousands of reversals. Certain adverse events were lower than with the 4 mg nasal spray, but participants expressed a desire for both injectable and nasal formulations. Given historical fluctuations in funding for and availability of naloxone, program participants would be best served knowing how to use both types of naloxone, and policies, standing orders, and laws should allow parity in access between different formulations. Differences in adverse event profiles between naloxone formulations [69,70] may be of relevance to policymakers. The behavior of titrating doses of naloxone to prevent adverse events also suggests that there may be underrecognized demand for formulations that deliver smaller or fractional doses, and reinforces the policy importance of ensuring that multiple forms of naloxone are available. Education on dose titration within harm reduction programs may also be an opportunity to prevent or reduce adverse events.

Fifth, sharing of naloxone between participants is a natural phenomenon, especially when provided at no cost to the participant. While secondary distribution of sterile syringes has been extensively studied in many countries [82–86], corresponding studies for naloxone distribution have not been published. Primary encounters with harm reduction staff provide opportunity for additional services to be offered (not just drug-related), a benefit that is attenuated through secondary distribution. Furthering this trend, over-the-counter versions of naloxone nasal sprays were approved by FDA in 2023, naloxone-dispensing vending machines are rapidly expanding [87], and mail order naloxone distribution has been established on a national scale [88], eschewing the direct human connection necessary for the comprehensive harm reduction services like Prevention Point Pittsburgh have traditionally provided. Concerted policy, technology, telehealth, and communication innovations could supplement these innovative distribution channels to re-establish more comprehensive care possibilities in an era where naloxone distribution is becoming more indirect. As demonstrated by the rich programmatic context in this paper, traditional harm reduction programs possess a depth of untapped applied experience that can inform broader policy and regulatory decisions.

### Future Research Needs

Severe precipitated withdrawal should not be dismissed as an “unavoidable” adverse event expected to occur in some OREs using antagonists, but rather needs to be studied independently, especially as it could lead to short- and long-term changes in behavior that increase risk for subsequent overdose; further research is needed to determine how withdrawal-related AEs can be reduced. For example, severe withdrawal can lead to immediate and repeated re-dosing with opioid agonists. In the weeks that follow an ORE, negative withdrawal experiences could also lead to use of street drugs alone. Co-author and Prevention Point Staff member MV shared testimony at a scientific conference articulating this connection. He had overdosed on heroin among strangers who administered 3 doses of naloxone, and he then received multiple unknown additional doses from uniformed first responders. He felt very anxious and was vomiting so frequently he had trouble breathing: “I tried to re-dose with heroin every 15 minutes to feel anything other than this horrible feeling. For months after that bad overdose, I was super hesitant to use around others. I mostly wanted to use alone to avoid something like that from happening again which put me at great risk.” [89]

To empirically establish post-overdose behavioral consequences, validated and easy-to-use outcome measures for overdose severity and response could be developed. Randomized field studies, such as comparing formulations of opioid antagonists, could also provide empirical evidence to inform policy and local purchasing decisions. Further research may also be warranted into other factors at the pharmaceutical formulation level, in terms of pharmacokinetics or packaging. And finally, training on timing and titration of dose administration and rescue breathing all bear scientific scrutiny. Qualitative studies involving those with lived experience should be considered to bring light to dimensions of community-based naloxone distribution and bystander naloxone administration that are not observable quantitatively [90,91]. Regardless of the method, there is pressing need for this type of work to more fully understand unmet needs in a changing environment.

## CONCLUSION

This comprehensive analysis of a harm reduction program reveals that while enabling state legislation can create the environment for expanded naloxone distribution, when naloxone is distributed to people not at risk of overdose or their immediate social networks, increases in dispensing volume can become negatively decoupled from actual administration and overdose reversal. Expanding services to underserved communities requires additional innovation. We also found that the long-term consistency of less than 2 doses per ORE, high survival rate, and robust utilization all lend confidence in prioritizing naloxone distribution directly to people who use drugs. Finally, we found lower rates of adverse events with lower doses of naloxone, titration, and with injectable intramuscular formulations. Collectively, these findings can help re-prioritize community-based naloxone distribution to those most likely to use the antidote to reverse an opioid overdose.

## Data Availability

Pre-registration: DOI osf.io/b2f4h. Codebook, data collection form, and analytic code: DOI osf.io/sq5d6. Data were provided by Prevention Point Pittsburgh and are used with permission. Due to the sensitive nature of syringe service provision, legal risks and stigmatization of drug use, and the risk of deductive disclosure, the decision was made to not make the data public. Interested parties can contact Alice Bell at Prevention Point Pittsburgh for collaboration and data requests.

## ACKNOWLEDGEMENTS

We thank the participants of Prevention Point Pittsburgh for their work in reversing opioid overdoses. We are indebted to Dan Bigg and staff at the Chicago Recovery Alliance who provided free naloxone to Prevention Point Pittsburgh in early years of this study and originated this intervention. We thank LaMonda Sykes, Bridgette Mountain, and Natalie Sutton for administrative support at UNC. We thank Alex Bennett and Tiffany Fitzpatrick for helping to implement naloxone distribution quickly in 2005 when overdose deaths began to spike from fentanyl in the opioid supply. Additional FDA project workgroup members included Mallika Mundkur, Tamra Meyer, Candice Collins, Chi-Ming (Alice) Tu, Celia Winchell, Bic Nguyen, Nushin Todd, Srikanth Nallani, Rigoberto Roca, Blair Coleman, Sanae Cherkaoui.

## TRIAL REGISTRATION

This investigation was pre-registered osf.io/b2f4h

## CODE AND DATA SHARING STATEMENT

Pre-registration: DOI osf.io/b2f4h. Codebook, data collection form, and analytic code: DOI osf.io/sq5d6. Data were provided by Prevention Point Pittsburgh and are used with permission. Due to the sensitive nature of syringe service provision, and the risk of deductive disclosure, the decision was made to not make the data public. Interested parties can contact Alice Bell at Prevention Point Pittsburgh for collaboration and data requests.

## CONTRIBUTORSHIP STATEMENT

ND, AB, MDS, EW, MN, AS, DC, ZD, and AM contributed to study conceptualization and design. AB and MV (and other Prevention Point Pittsburgh staff) conducted data collection and entry, and provided operational context. ND conducted the statistical analysis and is solely responsible for data integrity. All authors contributed to result interpretation and manuscript development.

